# Herpesvirus infections eliminate safeguards against breast cancer and its metastasis: comparable to hereditary breast cancers

**DOI:** 10.1101/2023.07.03.23292185

**Authors:** Bernard Friedenson

## Abstract

Breast cancer has no simple explanation. I tested the hypothesis that Epstein-Barr (EBV) infections promote the disease because they disable breast cancer safeguards. I used bioinformatics of public information from approximately 2100 breast cancers. Results demonstrate that chromosome breakpoints in breast and ovarian cancer cluster around the same breakpoints in diverse EBV-associated cancers. Cancers unrelated to EBV do not have these clusters. Breast cancers overexpress a methylation signature caused by active EBV infection. EBV remnants interspace between MHC genes and piRNA clusters as CRISPR-like evidence of past infection. I then found breast cancer breakpoints cluster around EBV docking sites. This clustering occurs because EBV breaks chromosomes and then damages breast cancers safeguards: BRCA1/2 pathways, SMC5/6, and mitotic controls. Finally, EBV causes the same losses that drive breast cancer metastasis. Activated EBV bypasses all these safeguards without large numbers of particles or continuing presence. Immunizing against EBV proteins may prevent breast, ovarian, and other cancers.

**Summary:** Human papilloma virus promotes cervical cancer because it disables tumor suppressors. EBV in breast cancer resembles this model. EBV variants disable a variety of molecular and cellular safeguards that protect against breast cancer.

## Introduction

At the time of breast cancer diagnosis, its causes are difficult to isolate from multiple risk factors. A human cancer virus is one such risk factor, but a tumor virus does not cause cancer by itself (*1*). For example, the human papillomavirus (HPV) promotes cervical cancer by inhibiting tumor suppressors p53 and Rb (*2*). Epstein-Barr virus /Human herpesvirus 4 (EBV/HHV4) is a risk factor for breast cancer since active infection increases risk by 4.75 to 6.29-fold (*3, 4*). Active EBV is significantly more prevalent in breast cancer tissues than in normal and benign controls (*5*). EBV is asymptomatic in most people, but the virus infects at least 90% of the population (*6*). EBV usually remains latent and does not express enough genes to trigger a host immune response. However, EBV does express latent gene products in sporadic and hereditary breast cancers (*3, 7, 8*).

Breast cancer prognosis depends on whether latent EBV infection activates into its replicative form (*9*). Activation brings massive changes to host chromatin methylation and structure (*10-12*). Breast cancers have hundreds of these changes (*13*). In breast epithelial cell models, once malignant transformation occurs, the virus is no longer required (*14*).

Despite this evidence, the role of EBV in breast cancer is still unclear. Fortunately, cancers in other tissues have proven associations with EBV infection and serve as bases for comparison. These cancers occur in epithelial tissue and in the hematopoietic system. Cancers associated with EBV include nasopharyngeal cancer (NPC), EBV-positive diffuse large B-cell lymphoma (DLBCL), and endemic Burkitt’s lymphoma (BL) (*15*). In NPC, 100% of malignant cells are EBV-positive (*16-18*). Nearly 8500 EBV variant forms were present in NPC patients. A single host had over 2100 variants, each differing only slightly from a reference viral genome (*18*). In the host, mutations interfere with innate immunity and constitutively activate an inflammatory response. Overexpressed NFKB is a hallmark of NPC, occurring in 90% of NPC (*19*).

NPC is an explicit model for EBV-linked cancer with many genomic similarities to breast cancer. Almost all stage III breast cancers overexpress NFKB (*20, 21*). Over 64% of NPC is deficient in a pathway that depends on the breast cancer susceptibility genes BRCA1 and BRCA2 (*19*). In 126 NPC patients from Greece and Romania, the most frequently mutated genes were BRCA1 and BRCA2 (55.5% and 33.3%, respectively). The most affected functions included homologous recombination repair, chromatin remodeling, and immune responses (*22*). EBV produces miRNAs that inhibit BRCA1 and consistently downregulate BRCA1 in primary NPC (*23*). EBV hijacks ATR and ATM signals that control BRCA1 and BRCA2 activity (*24*). EBV also uses the MRN endonuclease complex related to the same pathways at the virus origin of plasmid replication (*25*)

The BRCA proteins function in a pathway to accurately repair DNA crosslinks and breaks by homologous recombination. This sprawling, interconnected pathway includes Fanconi anemia (FA) gene products and is often identified as the FA-BRCA pathway. The FA-BRCA pathway is deficient in hereditary breast cancer. SMC5/6 complexes are also essential for FA-BRCA-mediated repairs (*26*). EBV infection destabilizes SMC5/6 complexes (*27-29*), so EBV interferes with the FA-BRCA pathway.

Like NPC, diffuse large B-cell lymphoma (DLBCL) is a known EBV-linked cancer and a second model to compare to breast cancer. DLBCL is the most aggressive and most common type of non-Hodgkin lymphoma. The best prognostic marker in DLBCL is homologous recombination status (the FA-BRCA pathway) (*30*). Many DLBCL patients have a long-standing, ongoing inflammatory process such as chronic lung, skin, or bone infections (*31*), and a defect arises in EBV immunity (*32*). The inflammatory response becomes deregulated and drives the lymphoma. Endemic BL is another potential breast cancer model with an explicit EBV infection and NPC-like chromosome structural changes (*15, 33, 34*). In DLBCL and endemic BL, EBV variant infection accompanies MYC translocations. These translocations replace normal MYC control with a highly active immunoglobulin regulator (*35-37*). All BL subtypes have this rearrangement, which drives the disease (*38*). Inappropriate c-Myc gene activation also occurs in DLBCL and NPC (*39*). Deregulated MYC is a well-known contributor to breast cancer, and normal MYC function depends on the breast cancer gene BRCA1 (*40*).

NPC, endemic BL (*34, 41*), and breast cancers (*42*) have chromosomes with too many centromeres. The duplicated centromere can lead to breakage, fusion, and bridging cycles. These cycles happen during cell division to destabilize the human genome (*43, 44*). Mitotic spindles pull chromatids with excess centromeres in too many directions, producing more chromosome breaks. Repair causes complex rearrangements and attracts APOBEC3 to create mutation foci (*42*). Viral factors and MHC gene losses support these events (*19*). The EBV protein BNRF1 initiates centrosome amplification in infected B-cells, but viruses lacking BNRF1 are less likely to cause this effect (*45*). Breast adenocarcinoma cells also have excess centrosomes that allow abnormal attachments to spindle fibers during mitosis (*46*).

EBV cancers have deficits like hereditary breast cancer with mutations in BRCA1 and BRCA2 genes. EBV activation may then be equivalent to inheriting this gene damage. Because this possibility predicts a role for EBV in breast cancer, I began this study by comparing chromosome breakpoints in known EBV cancers to breakpoints in breast and ovarian cancers. Landmark whole genome sequencing studies are invaluable and form the foundation for the project (*13, 42, 47-54*).

## Results

### Viral homologies around breakpoints in breast cancers from high-risk backgrounds cluster around breakpoints in NPC, an EBV-mediated cancer

EBV-mediated cancers such as NPC have defects in DNA repair and in inflammatory pathways, resembling hereditary breast and ovarian cancer. Because of this resemblance, I compared breakpoints in 70 NPCs to breakpoints in 139 breast cancer genomes from high-risk women (BRCA mutation, familial concentration, or young age). The results reveal that every chromosome in female breast cancers has breakpoints clustered near those in NPC.

Fig. 1A shows the exact distances between NPC and breast cancer breakpoints on chromosome 1 that are within 200,000 base pairs of each other. These nearest-neighbor breaks gather around a few low valleys that periodically occur across the whole chromosome. The exact distances between all matching breast cancer-NPC breakpoint pairs (including those further than 200,000 base pairs apart) give smeared results that are difficult to interpret (Fig. 1A). Different laboratories have collected this breakpoint data over many years. To allow for some differences, I split chromosome 1 into 5,000 base pair increments (2e-5 relative error) (Fig. 1B, supplementary TableS1). Breast cancer breakpoints cluster within 5,000 base pairs of NPC breakpoints, but the exact percentages are sensitive to the window size. Increasing the window to 175,000 base pairs (the length of EBV) substantially increases the agreement (see chr15, Fig. 1B, third row vs. lower right panel). The risk that any of these results is purely due to chance is less than 1 in 30,000, p<0.0001. Breast cancer and NPC breakpoint distributions are statistically the same for chromosomes 6,7,10,13,14,15,22, and X (TableS1). Breakpoints on some shorter chromosomes agree within 1000 base pairs.

**Fig. 1A.**
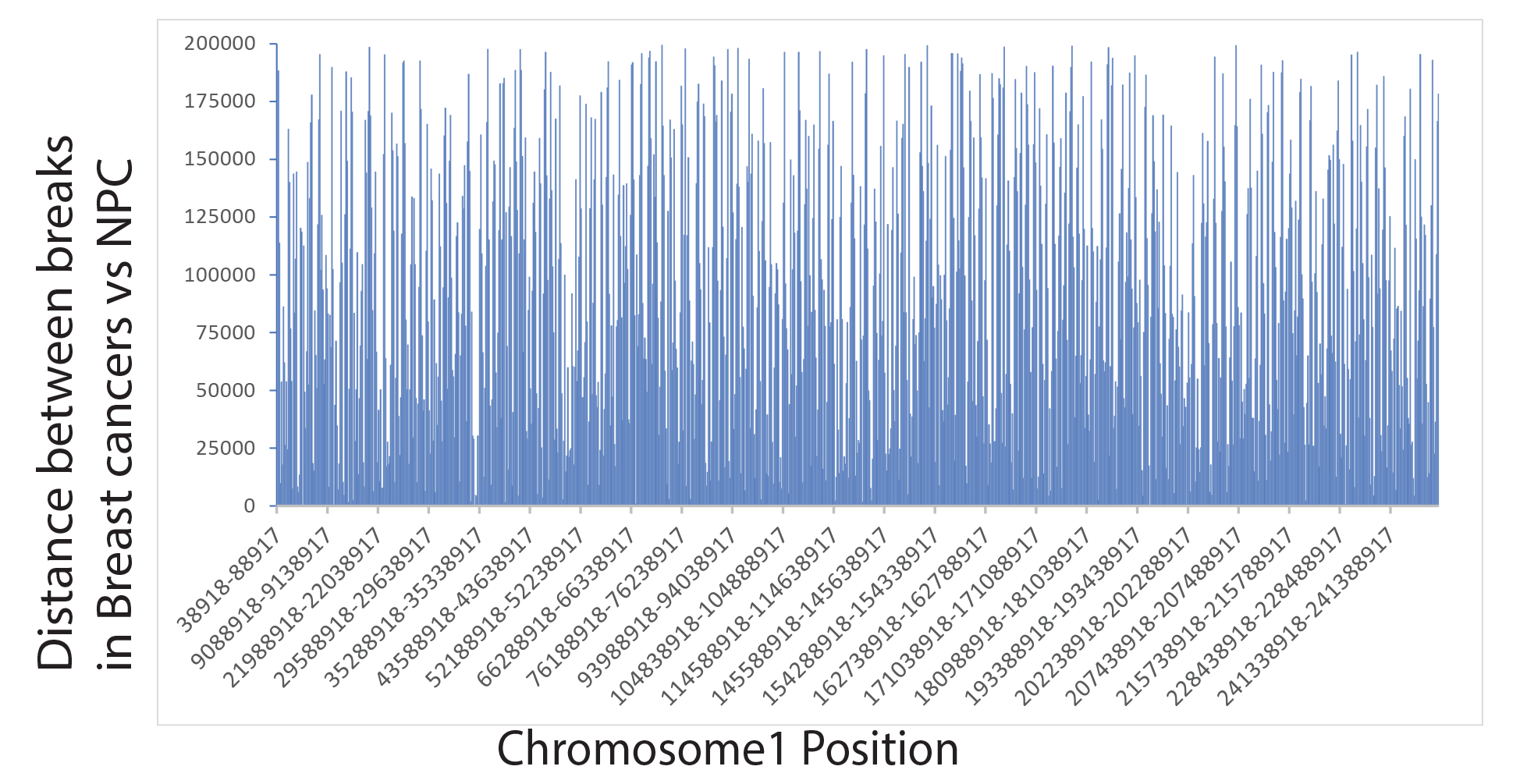
The exact distances between NPC and breast cancer breakpoints that are within 200,000 base pairs of each other.

**Fig. 1B.**
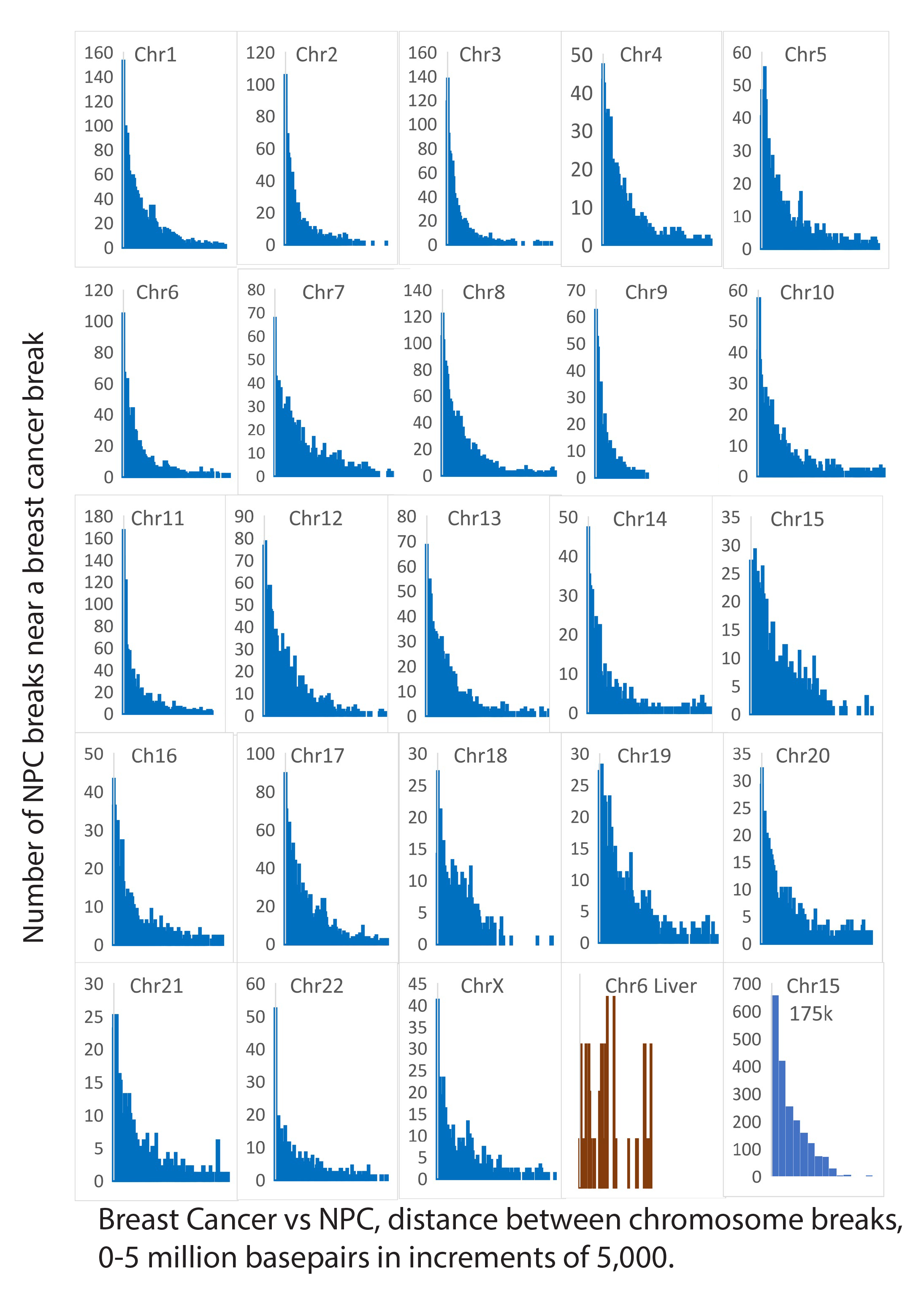
Breakpoints in 139 breast cancers from high-risk women (BRCA mutation, familial, or early onset) cluster around breakpoints in 70 NPC’s. On the X-axis, each bar on the graph is separated by 5,000 base pairs. The drawing at the lower right shows how selecting a larger bin size (the approximate length of EBV) affects Chr15. One example from an unrelated set of liver cancer data (lower right) does not show the same relationship to NPC. The panel at the lower right shows how the selection of a larger bin size (the approximate length of EBV) affects the distributions of breakpoints.

In contrast, cancers unrelated to EBV do not agree. Liver cancer breakpoints (*55*) differ from those in breast cancer or NPC (Fig. 1B, lower right). No breaks in 114 liver cancers on chromosome 1 are within 5,000 base pairs of breaks in any NPC; only one break on chromosome 6 in 61 liver cancers fits this window. According to a meta-analysis, the chance that breakpoints on chromosomes 1, 2, 6, and 8 were not within 5,000 base pairs in liver vs. NPC was 4.4 [CI=1.9-10]. NPC and liver cancers do not have the same breakpoint distributions (p=0.0004). DLBCL (likely EBV negative) had breakpoints far removed from those in NPC (Fig. 2G, discussed below).

The next step was to decide whether the similarity in chromosome breakpoint positions for breast cancers vs. NPC depends on BRCA1 or BRCA2 gene mutations. I compared breakpoint positions in high-risk women to those in sporadic breast cancers. The sporadic breast cancer group is 74 women, age >=70, with normal BRCA genes and no other known inherited cancer-associated mutations (*52*). Breakpoints these sporadic breast cancers cluster at chromosomal locations similar to the high-risk women, although frequencies and distributions differ significantly (Fig. 1C).

**Fig. 1C.**
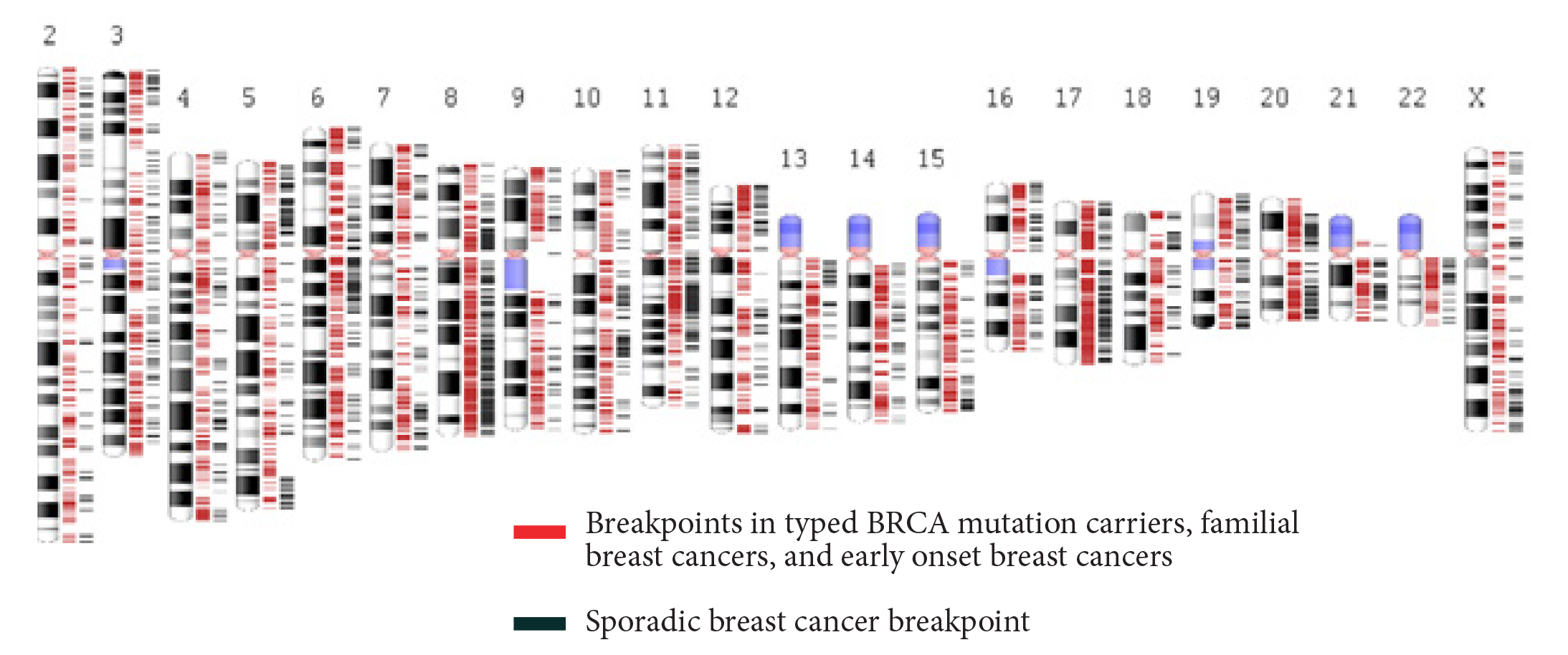
Inter-chromosome translocation break positions in 74 mutation-associated, familial, or early onset female breast cancers (red) vs. 74 likely sporadic female breast cancers (black).

I validated this conclusion in detail by comparing breakpoints in sporadic breast cancer to 70 NPCs. Breakpoints within 5,000 base pairs of an NPC breakpoint are the largest single category on most chromosomes (Fig. 1D). For example, chromosome 22 has 39 (33%) of its 118 breaks within <= 5,000 base pairs of an NPC breakpoint. According to Fisher’s test, chromosome 22 breakpoints in sporadic breast cancers and NPC are related (p<0.0001).

**Fig. 1D.**
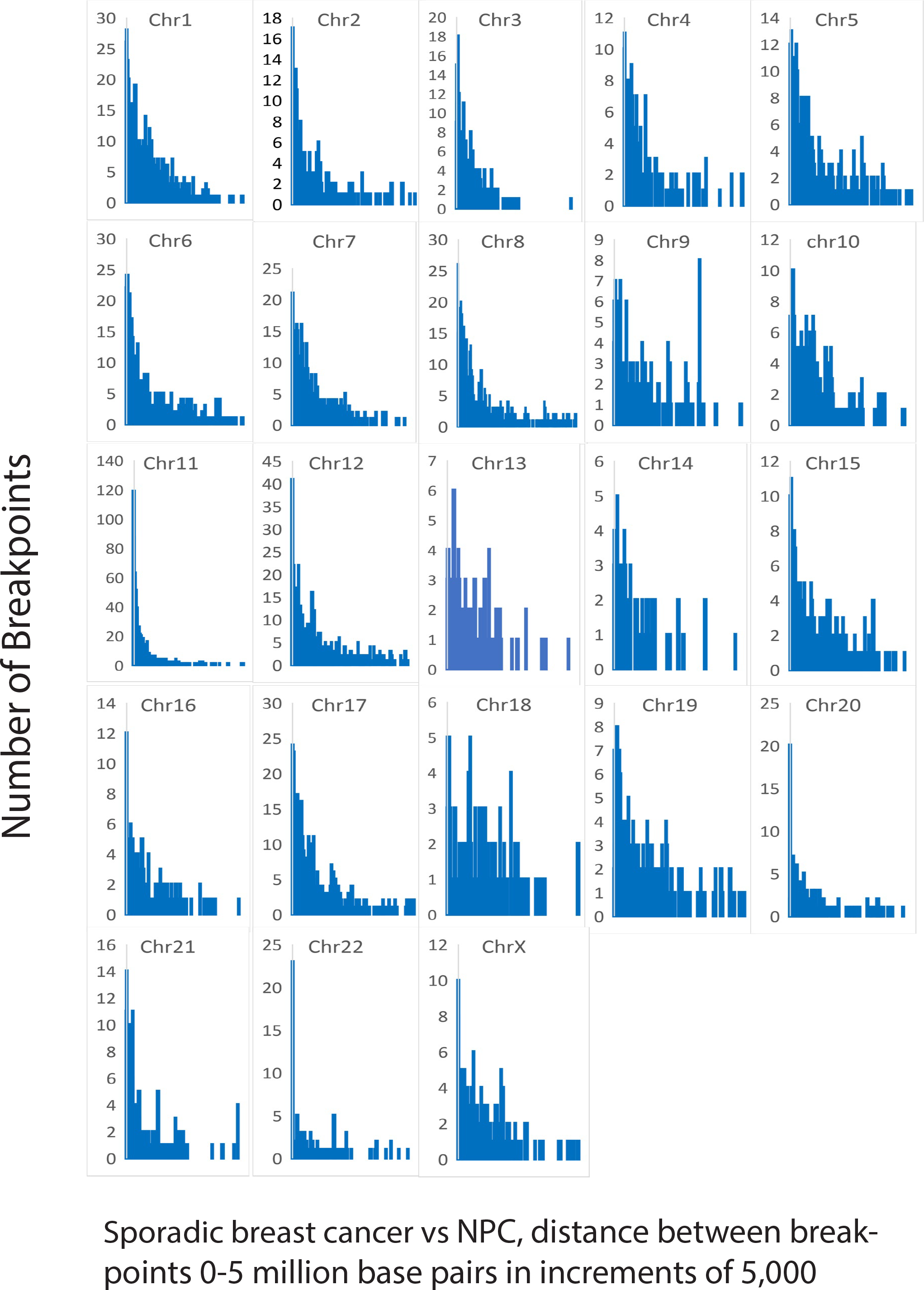
Breakpoints in 74 sporadic breast cancers cluster around the breakpoints found in 70 NPC’s. On the X-axis, each bar on the graph is separated by about 5,000 base pairs.

### Chromosome breaks in breast cancer subgroups

Next, I further tested the relationship between viral and breast cancers using data from breast cancer subgroups. Triple-negative breast cancers are likely to be BRCA1 positive (*56*), while HER2 amplification is uncommon in BRCA1 and BRCA2 mutation carriers (*57*). Fig. 1E shows breakpoints nearest NPC breakpoints vs. triple-negative and HER2-positive breast cancers (20 and 22 patients, respectively). Although subgroup differences are noticeable, breakpoints on all chromosomes are related to NPC.

**Fig. 1E.**
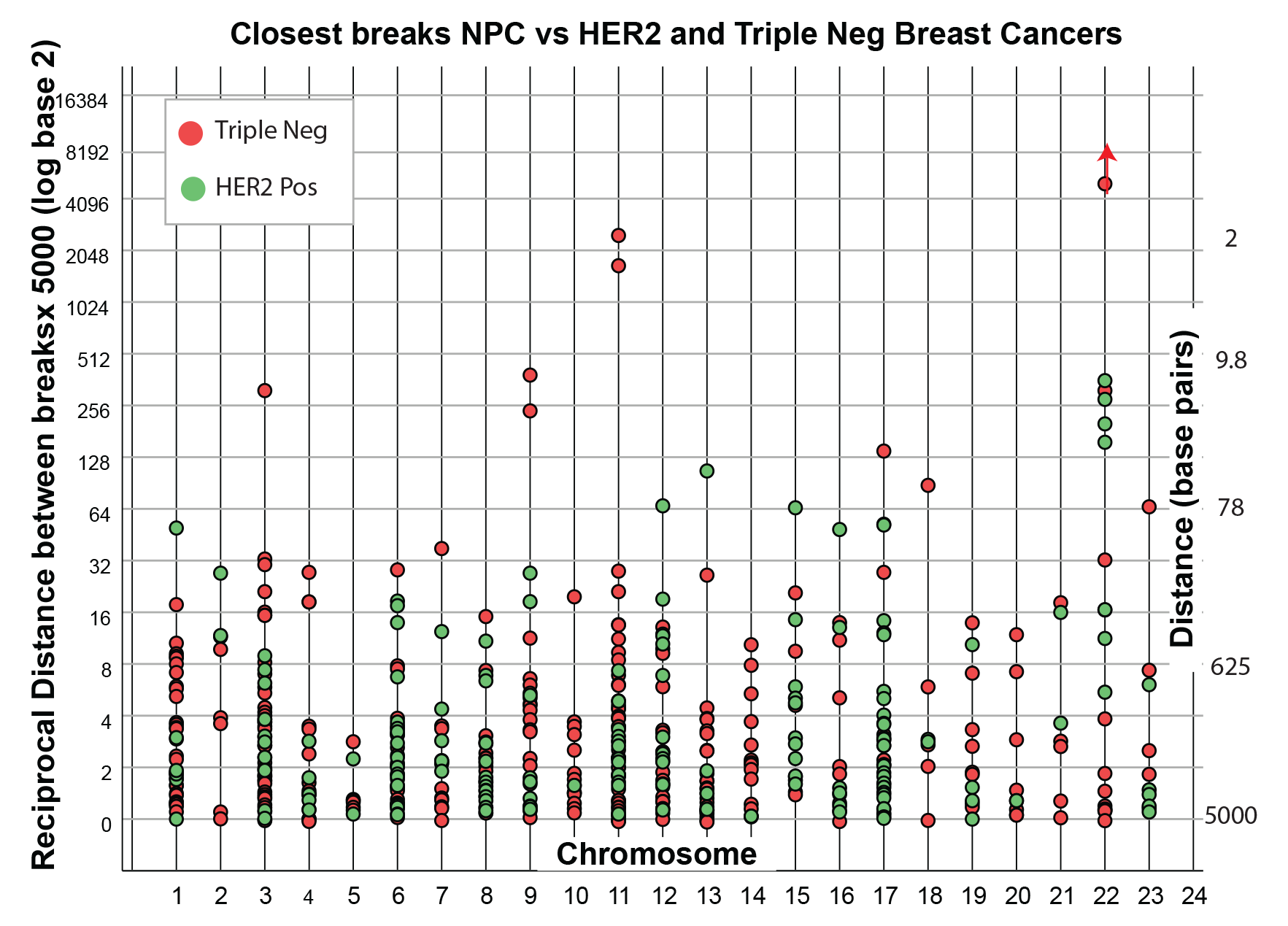
Breakpoints in BRCA-associated and non-associated ovarian cancers cluster around breakpoints in the 70 NPCs. The graph at the lower right represents chromosome 9 data remaining after removing all BRCA-associated ovarian cancers.

Four chromosomes, all essential for immunity, attract virus-like breakpoints in the two breast cancer subgroups. Chr3 includes code for multiple olfactory receptors, several genes needed to prevent cancer (MLH1, PIK3CA, VHL), and chemokine receptors required for inflammatory responses. Olfactory receptors are on macrophages, presumably permitting responses to toxic inhalation and controlling inflammation (*58*). HER2-positive breast cancers preferentially target Chr6. Chr6 includes the HLA antigen genes, a DNA protective mechanism (see below), and NFKB1L1, a presumptive NFKB inhibitor. A breakpoint hot spot around interferon receptor gene IFNGR1 compromises viral immunity. Chr11 has virus-like breaks in both HER2-positives and triple-negatives. Chr11 is home to the largest number of olfactory receptor genes. Chr17 encodes the TP53 DNA damage response gene and the BRCA1 gene.

Similar breakpoint distributions in NPC vs. high-risk vs. sporadic breast cancers vs. breast cancer subgroups argue that the causes are similar. The sporadic breast cancer patients are older than the high-risk women, arguing against age as responsible for similarity to NPC breakpoints.

### Viral homologies around breakpoints in mixed adenosquamous ovarian carcinoma also cluster around breakpoints in EBV-mediated cancer

The above results raise the question of whether similarity to the EBV cancer NPC is specific to the breast. Ovarian cancer is a logical test because BRCA1 or BRCA2 mutations can drive ovarian cancer, like breast cancer (*59*). Fig. 1F compares chromosome breakpoints in 24 mixed adenosquamous ovarian cancers to breakpoints in NPC. The results mirror breast cancer comparisons. (Figs. 1B, 1D, 1E). About half the ovarian cancers have likely hereditary BRCA mutations. A few test calculations removed these BRCA-mutated cancers, leaving sporadic ones. The sporadic cancers give the same results as the complete set but with fewer data (Fig. 1F, bottom right). These results show that breast cancer is not the only cancer with EBV-like breakpoint distributions. As in breast cancer, ovarian cancer breakpoint distributions cluster around NPC breakpoints without BRCA1 or BRCA2 gene mutation driver

**Fig. 1F.**
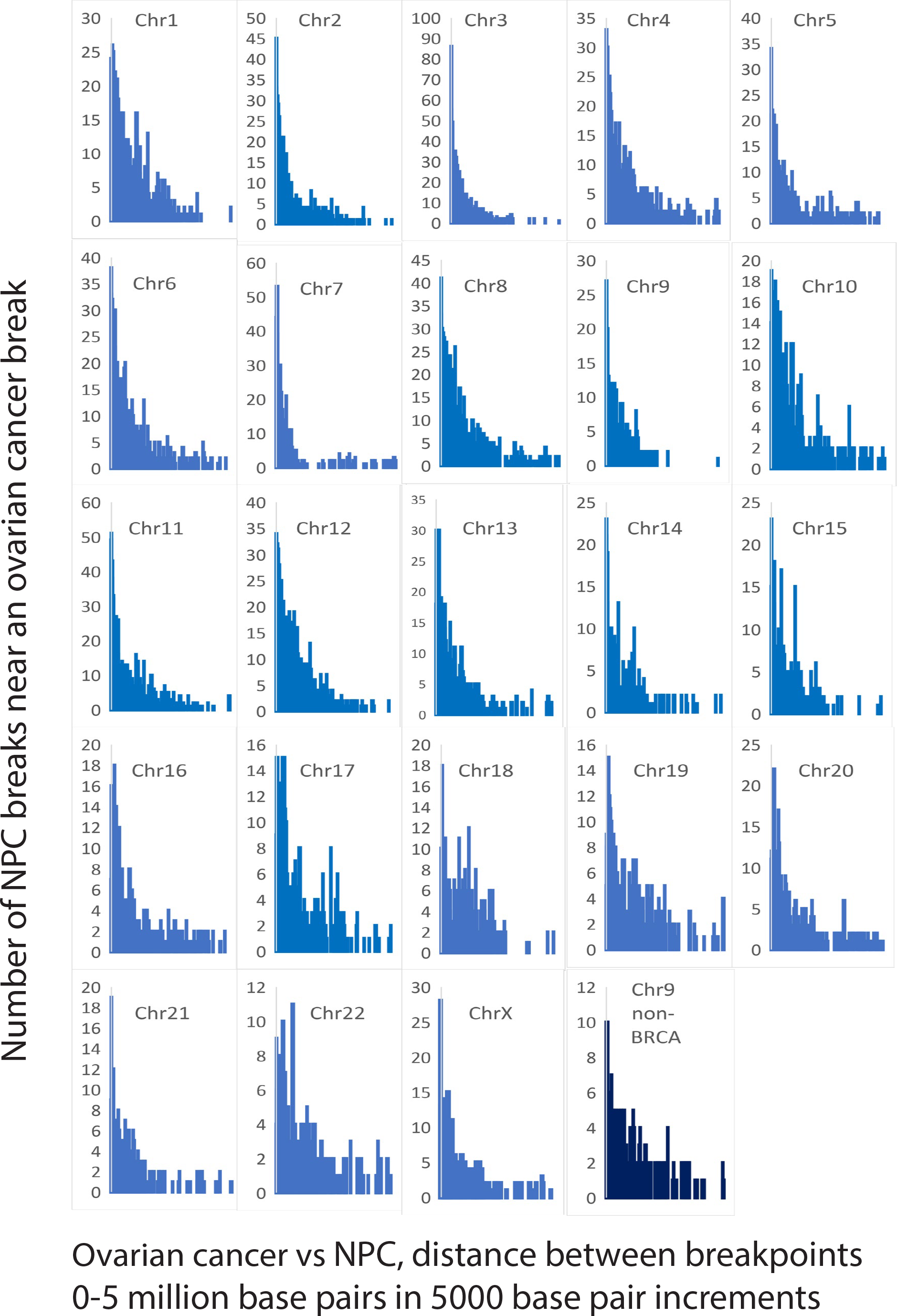
The log base2 of reciprocal distances between breakpoints in NPC vs. both triple-negative and HER2-positive breast cancers (20 and 22 patients, respectively). Higher points are closer.

### Breaks in lymphomas associated with EBV infection match breast cancer and NPC

EBV drives lymphomas as well as NPC. Based on epidemiologic research, FA-BRCA pathways protect against lymphomas (*60, 61*). If EBV is truly associated with breast cancer breakpoints, then breakpoint distributions in EBV mediated lymphomas should also resemble distributions in breast and ovarian cancers. Because MYC gene rearrangements drive some virus-associated lymphomas, the first step surveyed virus-like sequences surrounding the MYC gene. Fig. 2A shows MYC resides in a literal forest of retrovirus sequences (HIV1, FeLV, PERV, HERV) interspersed with EBV-like sequences. Breast cancer and lymphoma breakpoints disperse throughout the MYC region and beyond, but NPC breakpoints are less common.

**Fig. 2A.**
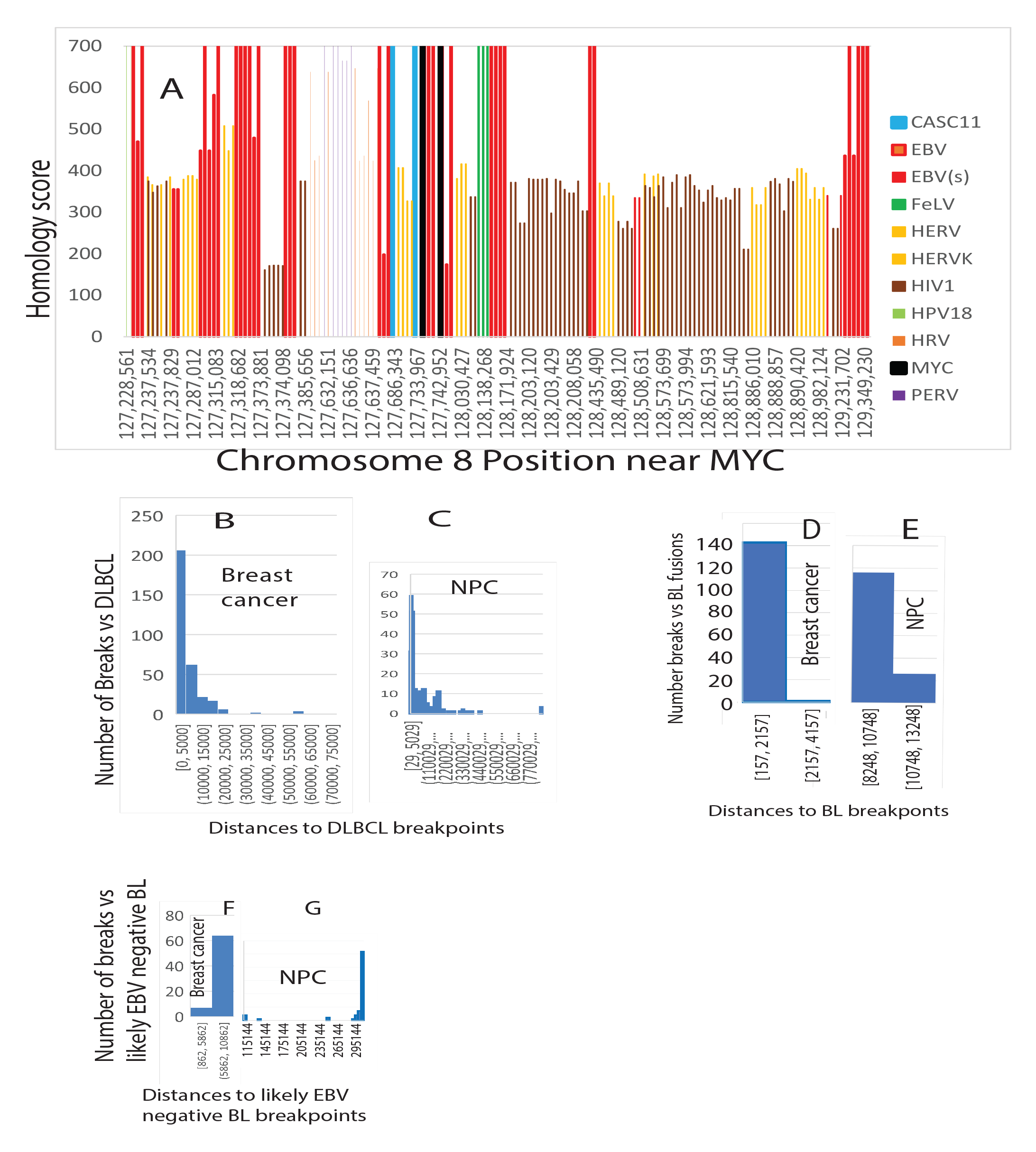
The DNA around the MYC gene on chromosome 8 is filled with virus-like sequences. CASC11 is an RNA gene that several cancers overexpress. Abbreviations: EBV(s), Stealth virus; FeLV, feline leukemia virus; HERV, human endogenous retrovirus; HERVK, human endogenous retrovirus K; HIV1, human immunodeficiency virus; HPV18, human papillomavirus 18, HRV, human retrovirus; PERV, porcine endogenous retrovirus.

The concentration of virus sequences around MYC on chromosome 8 prompted including the EBV-associated lymphoma DLBCL in breakpoint comparisons. The results find hundreds of breast cancer and NPC breakpoints congregate around breakpoint positions in 88 DLBCLs (*37*) (Figs. 2B, 2C). This agreement highlights multiple similarities among these cancers. A substantial percentage of DLBCLs have deficits in FA-BRCA-mediated homologous recombination repair (*62*). Some subtypes of DLBCL rely on NFKB activation (*63, 64*), which characteristically occurs in 90% of NPC (*19*) in response to EBV infection. About 79% of 47 EBV-positive DLBCLs have prominent NFKB activation (*32*). Inappropriate NFKB activation is common in breast cancer

EBV also drives a subset of BL, typically with MYC translocations. BL subsets can have mutations that impair homologous recombination (*65*), so BL breakpoint distributions should resemble the other cancers. An older set of BL cancers (*35*) has translocation breakpoints in the virus-rich area near the MYC gene, agreeing with >=140 breast cancer breakpoints (Fig. 2D). NPC does not have MYC gene rearrangements, with only a few breakpoints near MYC. Nonetheless, four different NPC breakpoints produce over 100 matches to BL translocation breakpoints beginning at about 8250 base pairs apart (Fig. 2E). An unpaired, 2-tailed t-test does not support a statistically significant difference between BL and NPC breakpoints in this area (p=0.69).

**Fig. 2B, C.** On chromosome 8, breakpoints in breast cancers (Fig. 2B) and NPC (Fig. 2C) cluster around breakpoints in DLBCL patients who were likely EBV-positive.

**Fig. 2D.** Based on MYC fusion sequences in BL ((*35*), breast cancer breakpoints on chromosome 8 cluster around BL breakpoints.

**Fig. 2E.** Even though NPC does not have MYC gene breakpoints, NPC breakpoints still cluster around BL fusion breakpoints, although at greater distances.

Sporadic BL does not involve EBV, so I used breakpoint data from sporadic BL as a control. This data represents a patient cohort tested for EBV with no positive test results (*66*). The sporadic BL patient breakpoints differ from EBV-driven BL (Figs. 2F and 2G). Less than half as many breast cancer breakpoints (Figs. 2F vs. 2D) are twice as far away from the control sporadic BL breakpoints. NPC has no relationship to the sporadic BL control (Fig. 2G), with hundreds of thousands of base pairs separating breakpoints in the two cancers.

**Fig. 2F.** For likely EBV-negative BL, half as many breast cancer breakpoints cluster around BL breaks at twice the distance.

**Fig. 2G.** For likely EBV-negative BL, NPC breakpoints are distant from BL breakpoints.

### Breakpoints occur near human sequences that resemble viruses in all breast cancers tested

The next step was to better understand the prevalence of viral relationships to chromosome breakpoints. I extended comparisons of human breast cancers in high-risk women (*52, 54*) to whole chromosomes. Virtually every breast cancer likely has breakpoints near sequences homologous to EBV. For example, chromosome 8 alone has 59,566 significant viral homology scores (>200), representing 128 breast cancer patients and 43,491 unique breakpoints. Breakpoints in 123/128 breast cancers were within 10,000 base pairs of a virus sequence. In 106 patients, the virus was an EBV variant with 3086 matching human sequences. According to Fisher’s exact test, chromosome 8 breakpoints and EBV variant sequence matches are not independent (p<0.0001).

Maximum homology scores over 4000 for human DNA vs. herpes viral DNA are abundant on all chromosomes tested. The 4000 score is 97% human-virus identity over nearly 2500 base pairs, with E "expect" values (essentially p-values) equal to 0. HKNPC60 and HKHD40 were virtually identical to human breast cancer DNA at many positions throughout chromosome 8 (145,138,636 base pairs) and chromosome 17 (83,257,441 base pairs)(Fig. 3A).

**Fig. 3A.**
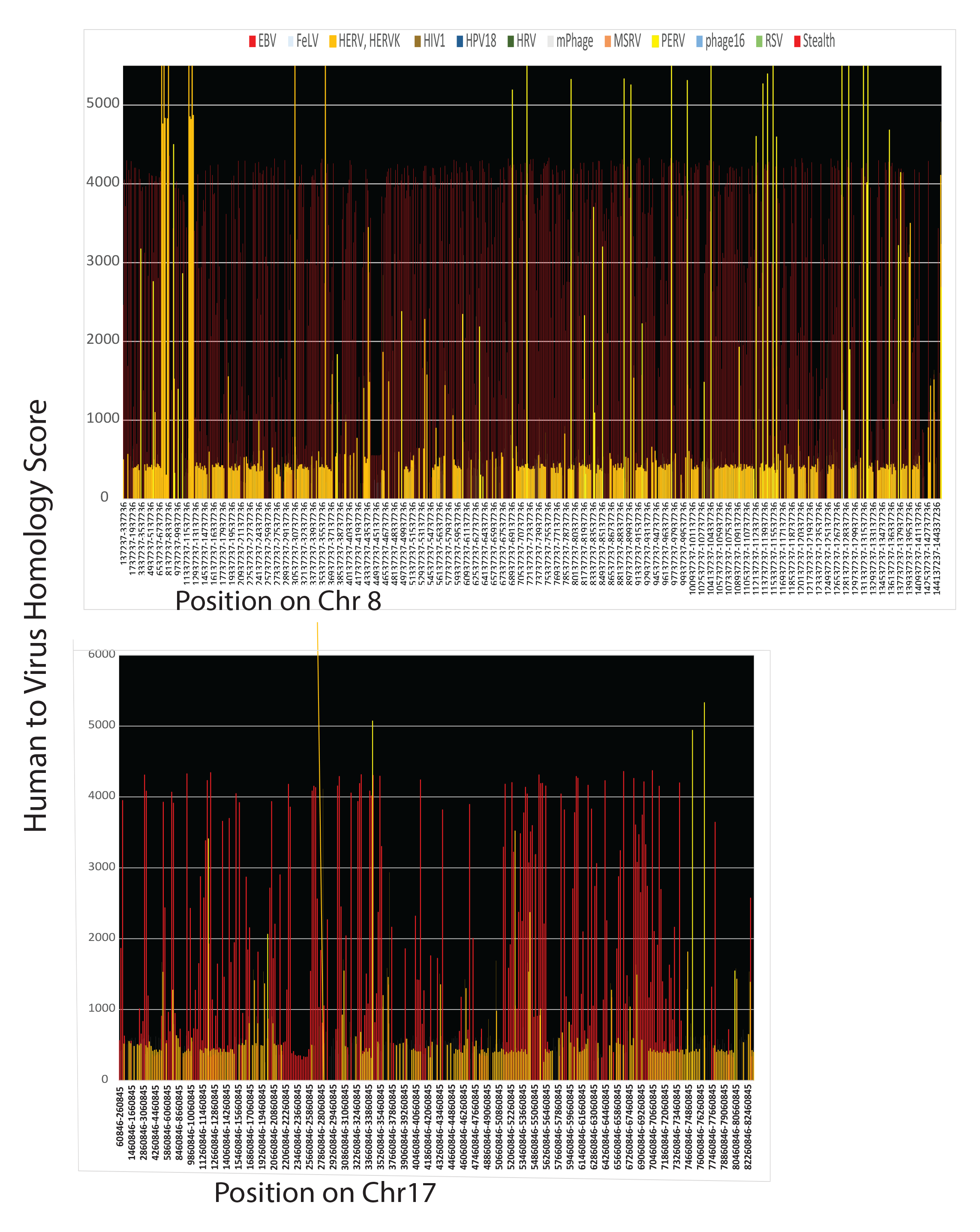
All viral homologies on the entire lengths of chromosome 8 (top) and chromosome 17 (bottom).

Viral sequences are not uniformly distributed among human chromosomes. On chromosome 8, the EBV variants HKNPC60 and HKHD40 had ∼11,000 human counterparts interspersed with 6,000 matches to endogenous retroviruses. Chromosome 17 (Fig.3A, lower panel) reverses the relative numbers of human positions that match EBV (1462) vs. endogenous retroviruses (10,445). Sequences near the HER2 gene are enriched in retroviral sequences (see below).

HKHD40 and HKNPC60 kept appearing in comparisons to human sequences, so I tested whether these variants were unusual. Fig. 3B shows hundreds of human gamma herpesvirus 4 variants almost identical to HKHD40 and HKNPC60 over at least 2,000 base pairs. The matching sets of viruses include many high-risk herpesvirus isolates from NPCs (*67*). Based on this information, HKHD40 and HKNPC60 strongly resemble other herpesvirus isolates, including many that confer high risks for NPC (*18*).

**Fig. 3B.**
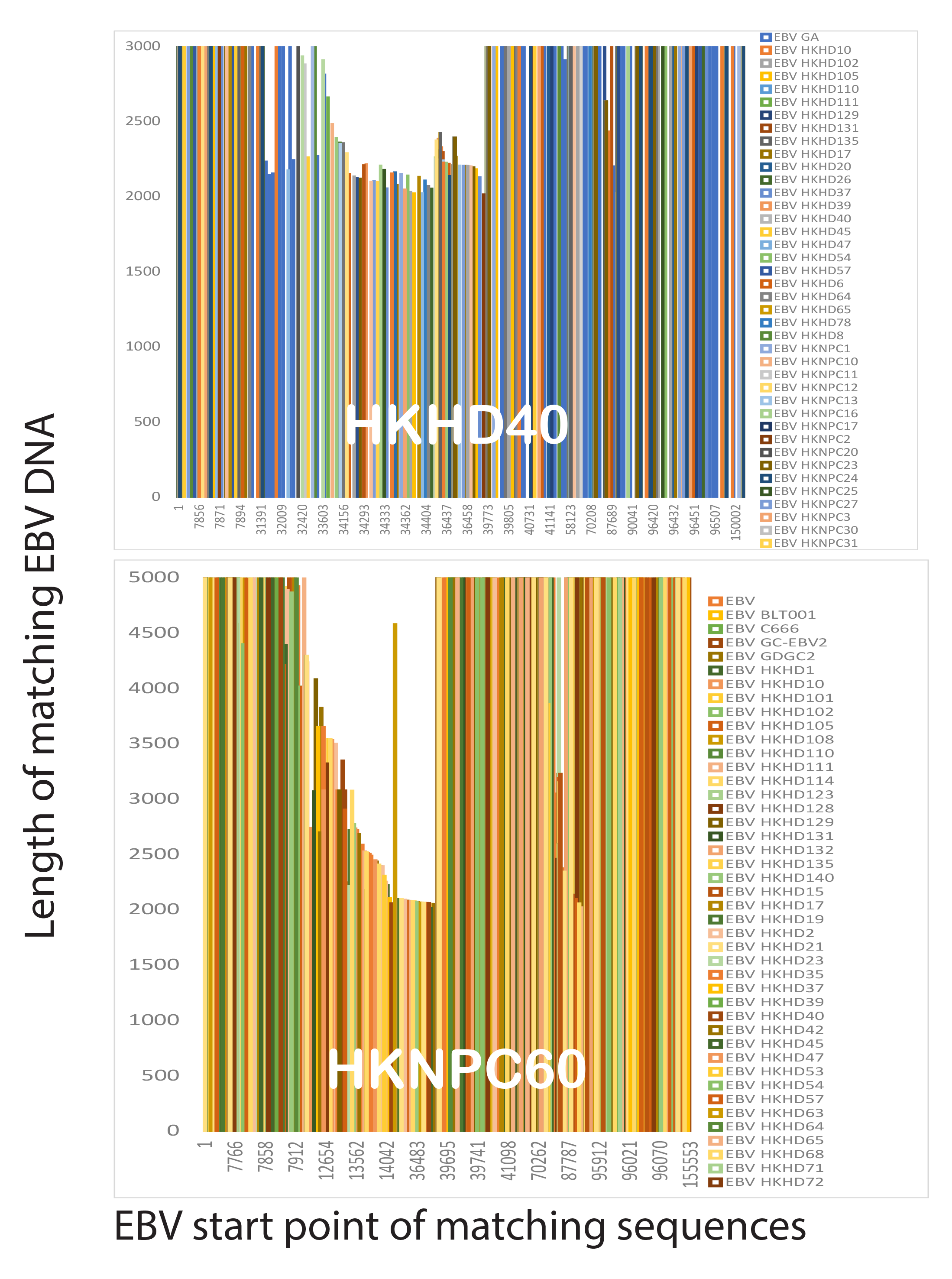
EBV variants HKHD40 and HKNPC60 are typical of hundreds of other EBV variants.

### Evidence of EBV infection

The evidence thus far supports the central hypothesis that EBV accompanies breast cancer because activated EBV disables tumor suppressor mechanisms. A significant obstacle is that disabling tumor suppressors does not require large numbers of viral particles or continuing virus infection; with so little virus, how can there be convincing evidence that EBV activates in breast cancer? Unlike retroviruses, EBV and its variants do not have integrase enzymes, so EBV has no conventional way to insert itself into the human genome. EBV integrates very rarely, with only one or two copies in BL cell lines (*68*). Using BLAST analysis, I tested EBV sequences against the human genome and found about 65,000 areas of strong homology (E<<e-10). Because 65,000 is far more than realistic EBV integration events, I wondered whether some EBV sequences were fragments created by a human version of the bacterial CRISPR system.

A candidate for such a human CRISPR version exists on chromosome 6p21.3, which includes MHC class I, II, and III genes. HLA antigens HLA-A, HLA-B, HLA-C, and HLQ-DMB are examples. HLA antigen variants are strong risk factors for NPC infections (*69*) because the HLAs break down antigens and display pieces of them to the immune system. Thirteen breast cancers listed in COSMIC have a deletion near this HLA region. About 23% of breast cancers have mutations directly affecting HLA class I or II genes. Many more breast cancers have indirect connections with multiple damaged genes that interact with HLA antigens or are otherwise essential for immunity. The region also holds NFKB1L1, a negative regulator of the NPC hallmark, NFKB. The 139 breast cancers from high-risk women have 284 breakpoints at chromosome 6p21.3. Breakpoints in the 70 NPC cancers also cluster there, with 40 breakpoints between Chr6:27,865,296-34,017,013.

In general, the piRNA system loosely resembles the bacterial CRISPR/Cas system, so I compared the distribution of piRNAs to the distribution of viral fragments in the MHC region of chromosome 6. The piRNA system inactivates virus-derived transposons (related to human endogenous retroviruses) by methylating or cleaving them. Fig. 4A shows a striking similarity in how remnants of exogenous and endogenous viruses distribute at 6p21.3. Remnants of both virus types are homologous to the same human sequence, and both types sandwich between piRNA sequences, sometimes right next to each other. Most sandwiches are at a regular interval or multiple (Fig. 4A). The sandwich arrangements are easier to see in Fig. 4A (bottom), which shows an arbitrarily selected 5 million base pair segment in this area of chromosome 6.

**Fig. 4A.**
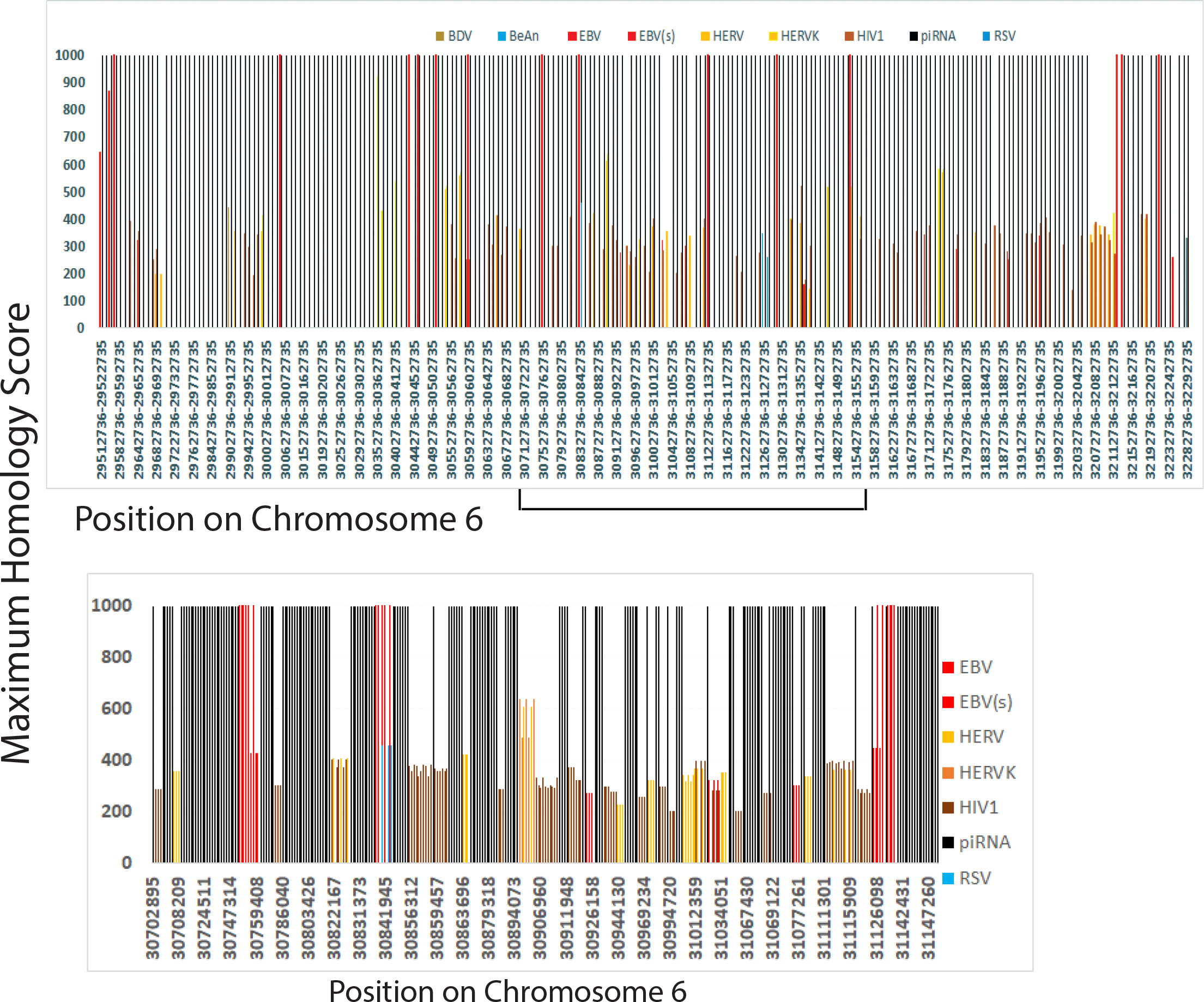
Distribution of remnants of exogenous and endogenous viruses over 2.77 million bases on chromosome 6p21.3. Smaller regions of interest show more detail.

This interspaced arrangement looks so much like CRISPR that it suggests piRNA defense mechanisms have inactivated EBV variants in addition to endogenous viruses. Long stretches of endogenous transposon-like DNA sequences routinely match exogenous viruses. Fig. 4B demonstrates that the same human DNA interval has homology to endogenous transposons (HERV) and exogenous viral sequences (HIV1, Stealth 1, BeAn58058, HPV16, and Zika). The piRNA system can store the same piece of DNA to protect DNA against these different viruses. HERV sequences are more abundant than exogenous viruses, explaining why I found intervals where HERV transposons are the only virus-like sequences present.

**Fig. 4B.**
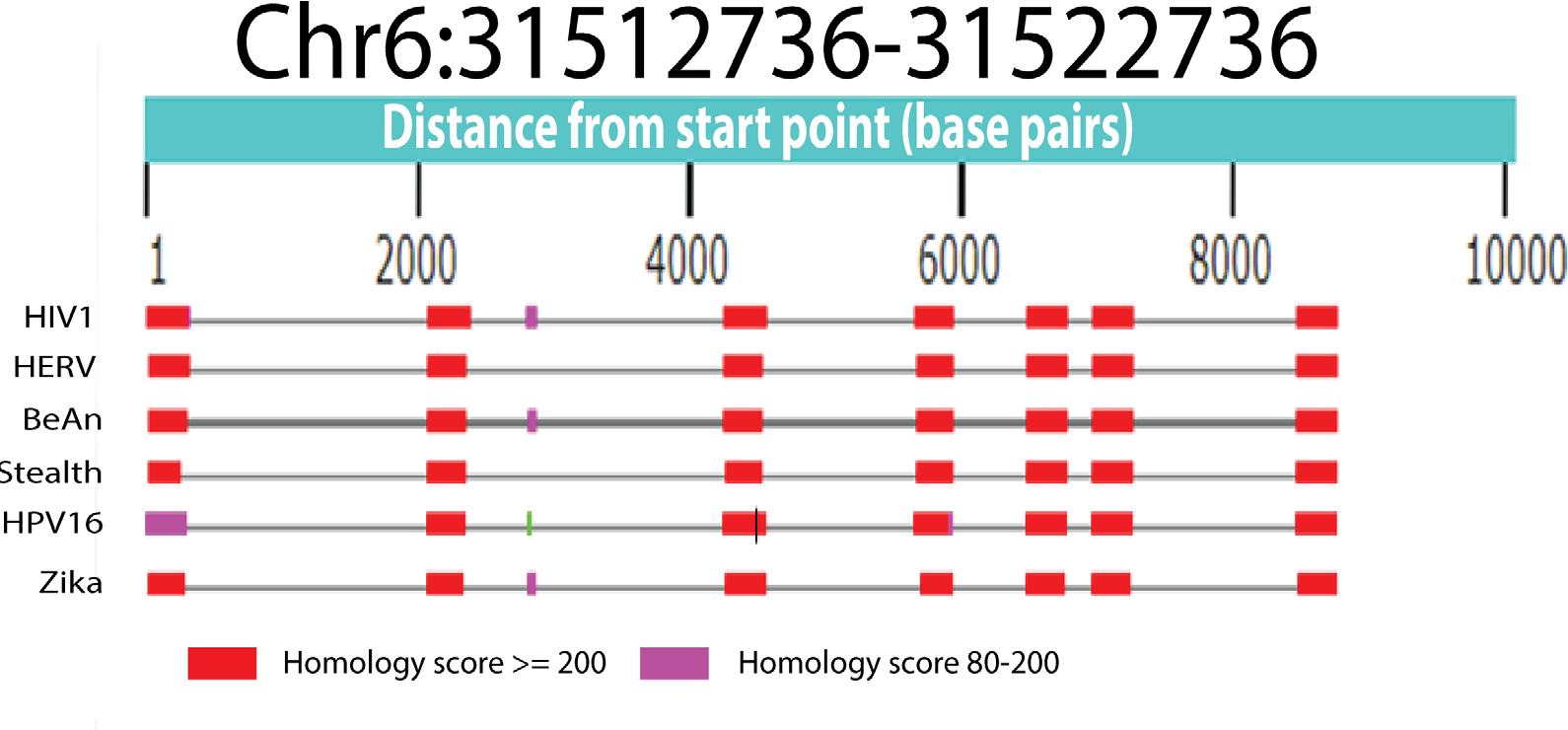
Transposons as endogenous retrovirus fragments are not easily distinguished from exogenous viruses because incorporated fragments strongly resemble each other.

The levels of various piRNAs vary by more than 1,000-fold, but the most abundant piRNAs are the only ones present in every cell. These abundant sequences drive the inactivation of foreign DNA. Rare piRNAs do not function in every cell but can potentially adapt to new genome invaders (*70*). The human genome organizes piRNA sequences into clusters (*71*). PiRNA sequences cluster near the MHC region of chromosome 6 (at ∼30-40 Megabases), with hundreds of piRNAs nearby (Fig. 4C).

**Fig. 4C.**
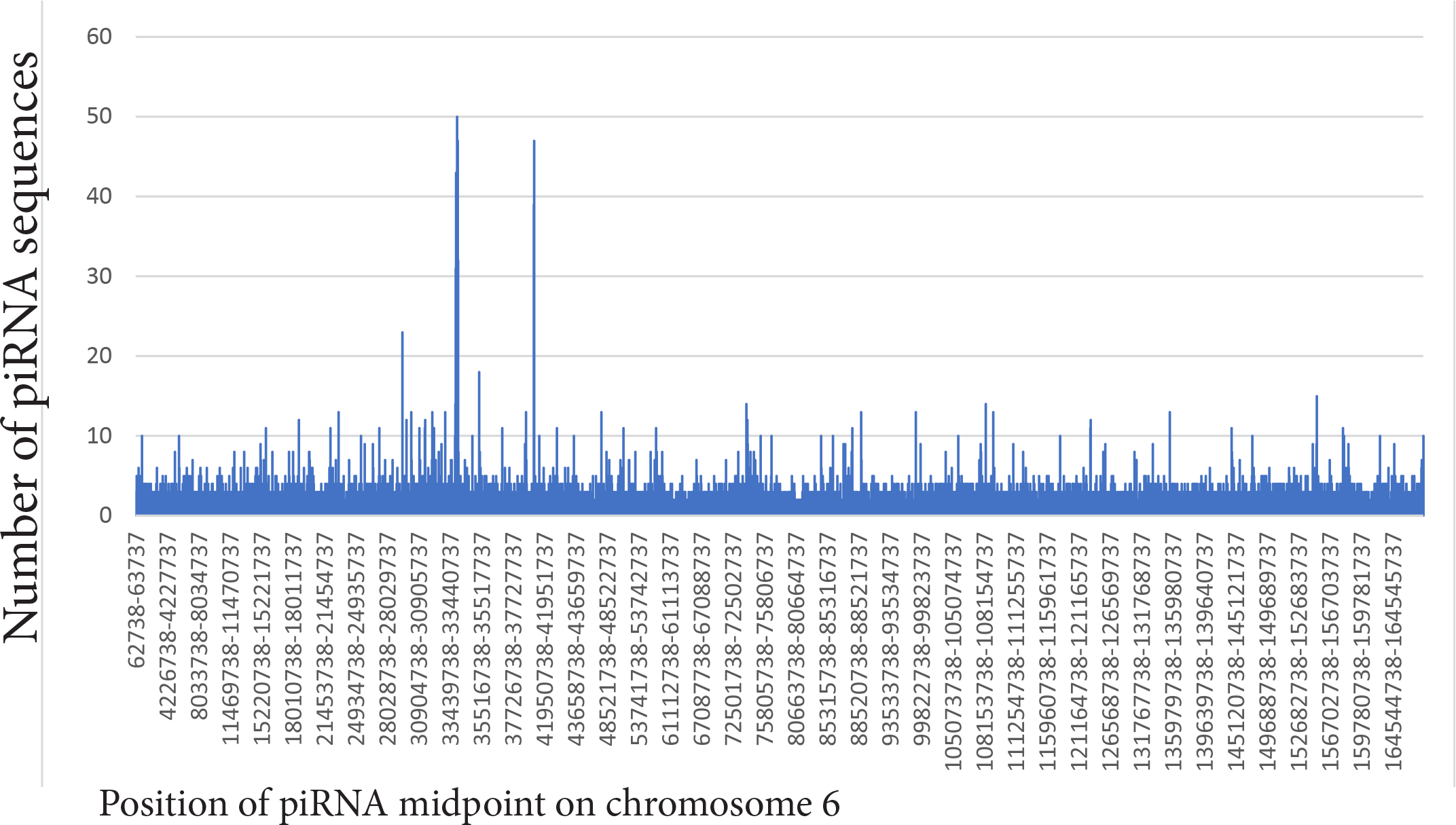
piRNA sequences are abundant near chromosome 6p21.3.

I obtained further evidence for EBV infection in breast cancer because I found its likely signature. Chromosome 6p21.3 contains an EBV infection marker (*72*). Using existing methylation data (*13*), I examined the marker in 1538 breast cancers. Fig. 4D shows that promoter methylation in this marker region differs significantly from normal controls. Fig. 4D shows hypermethylation of STK19, an MHC class III region gene (*73*) for RNA surveillance (*74*). In Fig. 4D, hypermethylation also inhibits a gene for preventing tumors (TNFB)(*75*) and a gene for responding to antigen-antibody complexes (C2). Polymorphisms in HLA-DMB antigen and SAPCD1, another class III MHC gene (*76*), at 6p21.3 have links to Kaposi’s sarcoma (*77*). HHV8 (KSHV) is a Kaposi’s sarcoma driver and is closely related to EBV (see below).

**Fig. 4D.**
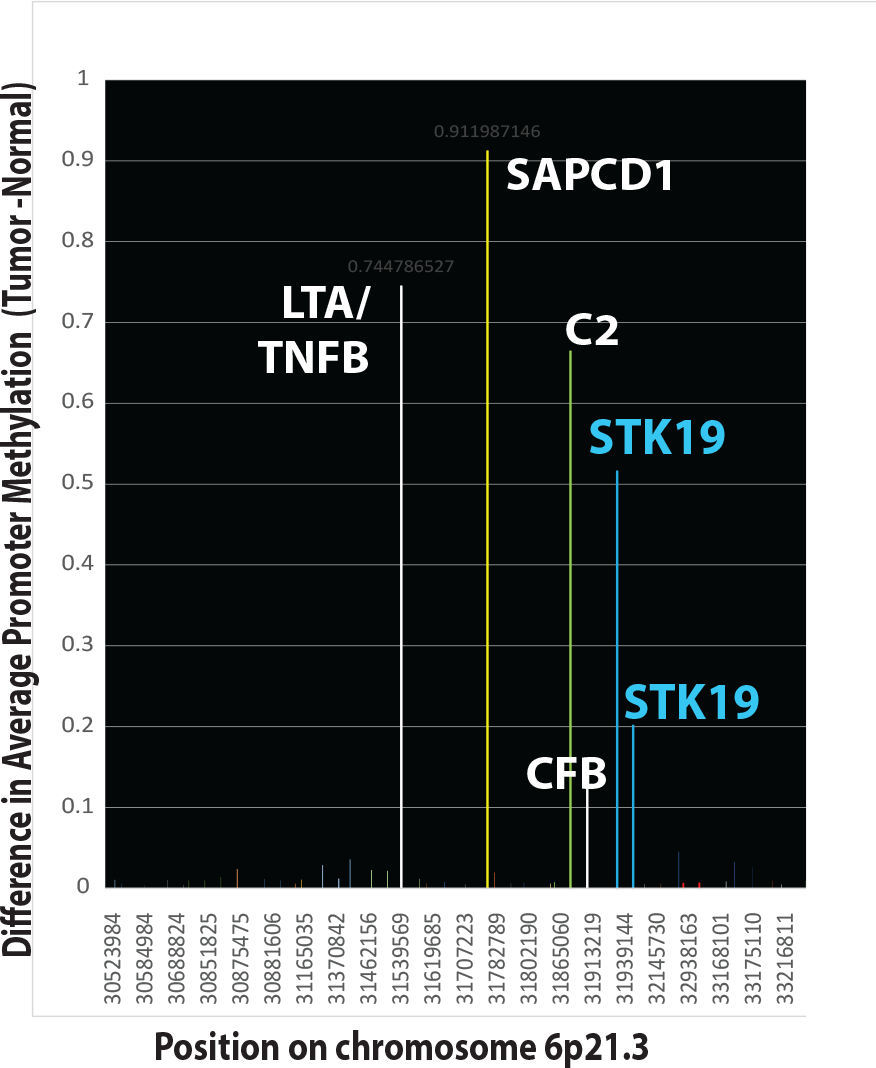
Chromosome 6p21.3 also contains a potential EBV infection signature (*72*).

Comparisons of viral sequences and piRNAs at 50-base pair resolution reveal more nuanced details of their arrangement (Fig. 4E). A few short EBV variant sequences fit within the borders of two piRNAs (darker red areas). Longer EBV variant sequences encompass one or two piRNAs, suggesting that inactive viral fragments can contribute piRNAs.

**Fig. 4E.**
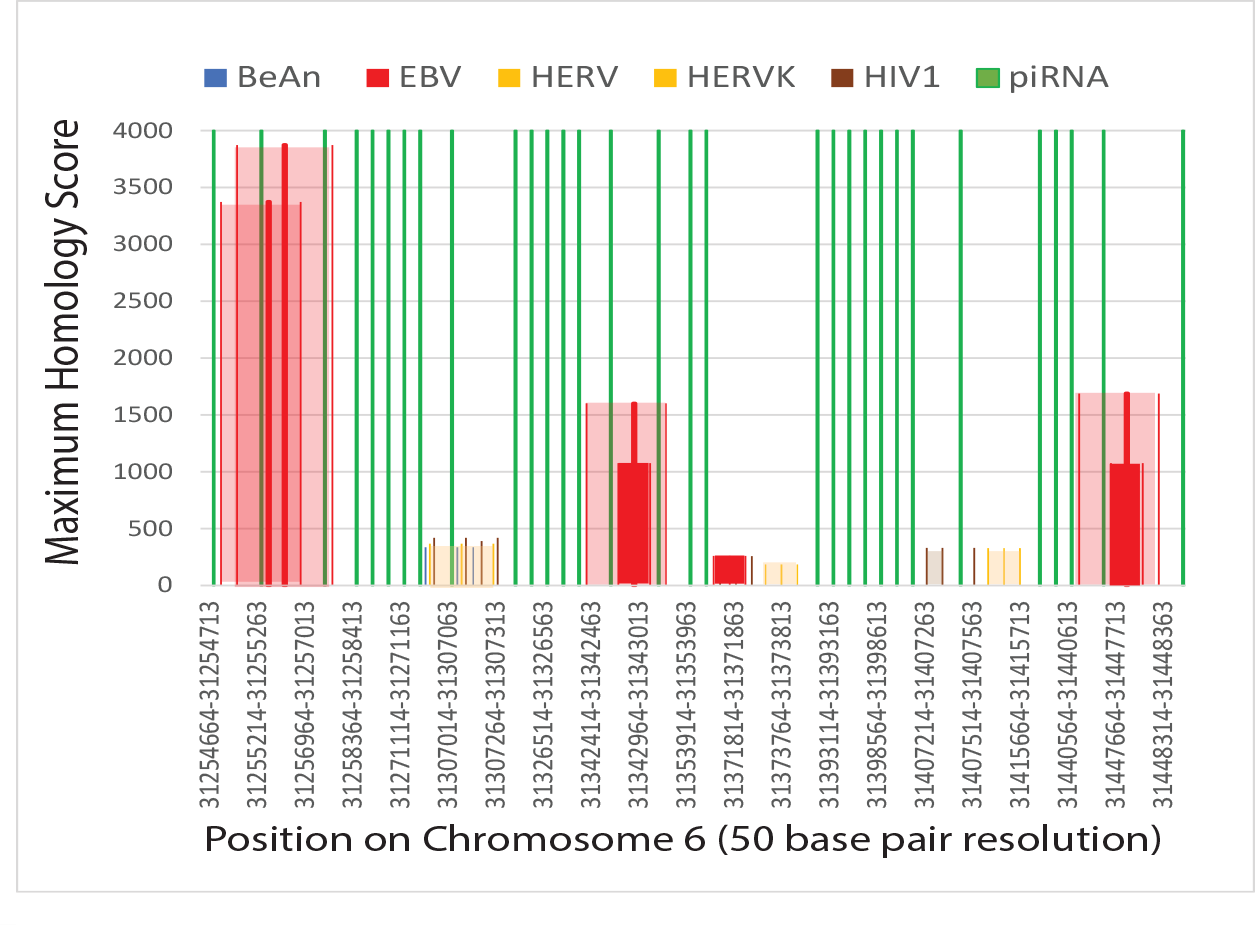
High-resolution 50 base pair comparison positions of piRNAs midpoints to virus-like intervals on chromosome 6. Each virus match is plotted as start-midpoint-end. The entire human chromosome region encompassed by viral homology is shaded.

In contrast to the clustered piRNA sequences on chromosome 6, other chromosomes have comparatively few piRNA sequences. For example, chromosome 17 contains piRNA clusters (*71*), but a virus-rich region of chromosome 17 at 50.00-56.47 million bases has few piRNAs. EBV variants are not always confined to large groups of piRNA sequences.

### Genes at the most frequent EBV tethering sites cluster around breast cancer breakpoints

In previous sections, breast and ovarian cancer breakpoints distribute most frequently near a characteristic set of breakpoints associated with EBV activation. Cancers unrelated to EBV do not match this set of breakpoints. Does EBV cause breast cancer breakpoints?

EBNA1 crosslinks viral and human DNA in the nucleus where viral episomes dock to human DNA. I found one EBV docking site on chr11 near FAM-D and FAM-B genes (*78*), close to two breast cancer breakpoints (Fig. 5A). This chr11 site is very near EBNA1 binding at strings of imperfect 18-base pair palindromes (chr11:114,604,212-114,625,620). This single locus is a significant EBNA1 binding site with tandem arrays of palindromes that can differ in 8 of 18 positions (*79*) (variants at six places and two mismatches). To test whether this was the only locus related to EBNA1 binding and breast cancer chromosome breakpoints, I looked for breast cancer breakpoints near other EBV docking sites. Some of the best information on EBV docking sites comes from 4C-chromatin capture experiments in EBV-positive BL cells. The largest numbers of breast cancer breakpoints on most chromosomes cluster around the genes (*10*) nearest to EBV tethering sites (Fig. 5B). BL cells providing the data have up to 1569 EBV docking sites distributed on all chromosomes (*10*), not just chromosome 11. EBV docking sites on chromosome 11 near the LUZR2 and FAT3 genes in BL cells (Fig. 5B) are more than 100 million and 21 million base pairs, respectively, from the 18 bp imperfect palindrome interval. Graphical estimation of virus tethering sites on chromosome 2 from these EBV-positive cells agree with breast cancer breakpoints (Fig. 5B, lower right).

**Fig. 5A.**
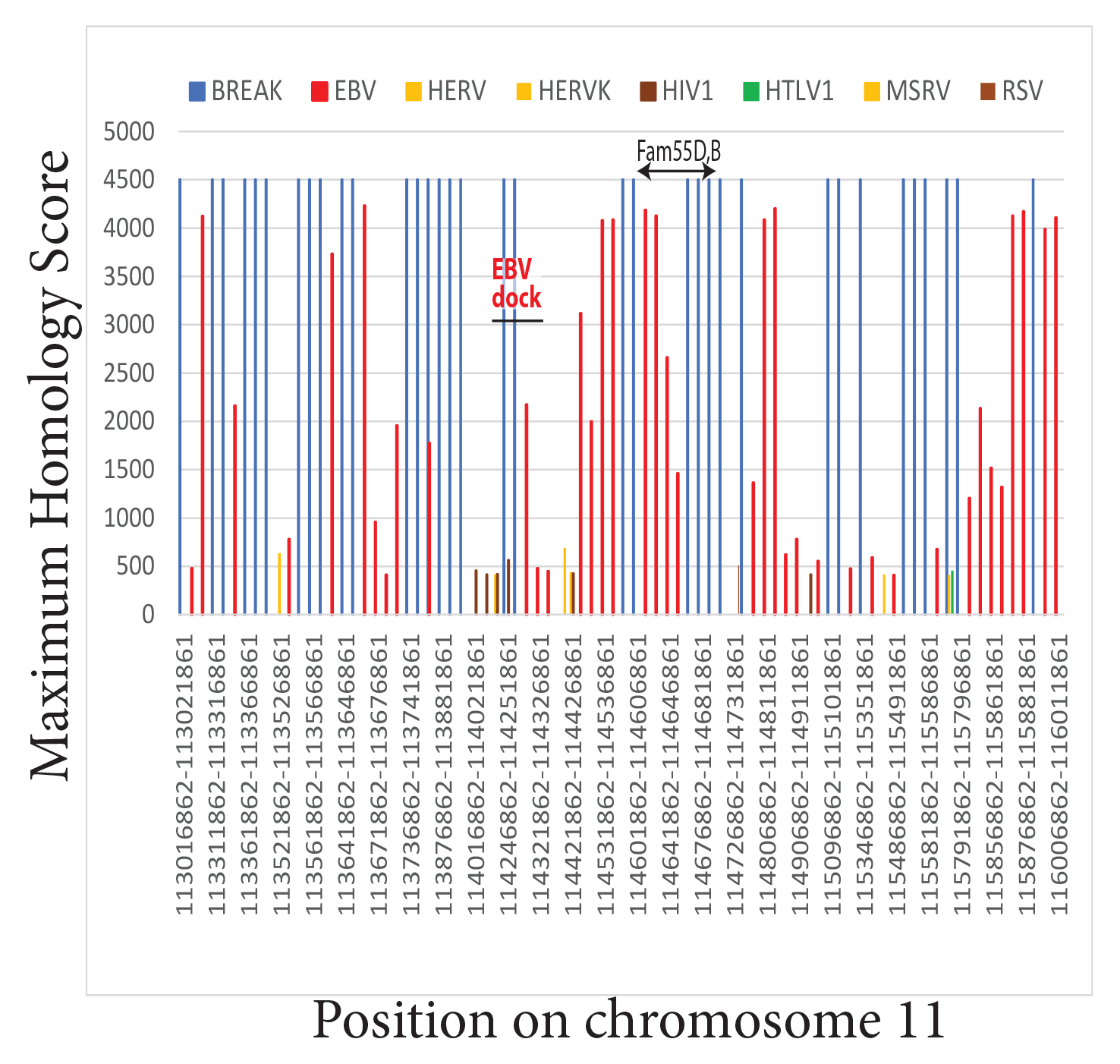
Maximum homology to human DNA for all viruses (y-axis) is plotted for known EBV genome anchor sites on chromosome 11 near FAM55 gene coordinates vs. breast cancer breakpoints. Based on data from Lu et al. ((*78*)

**Fig. 5B.**
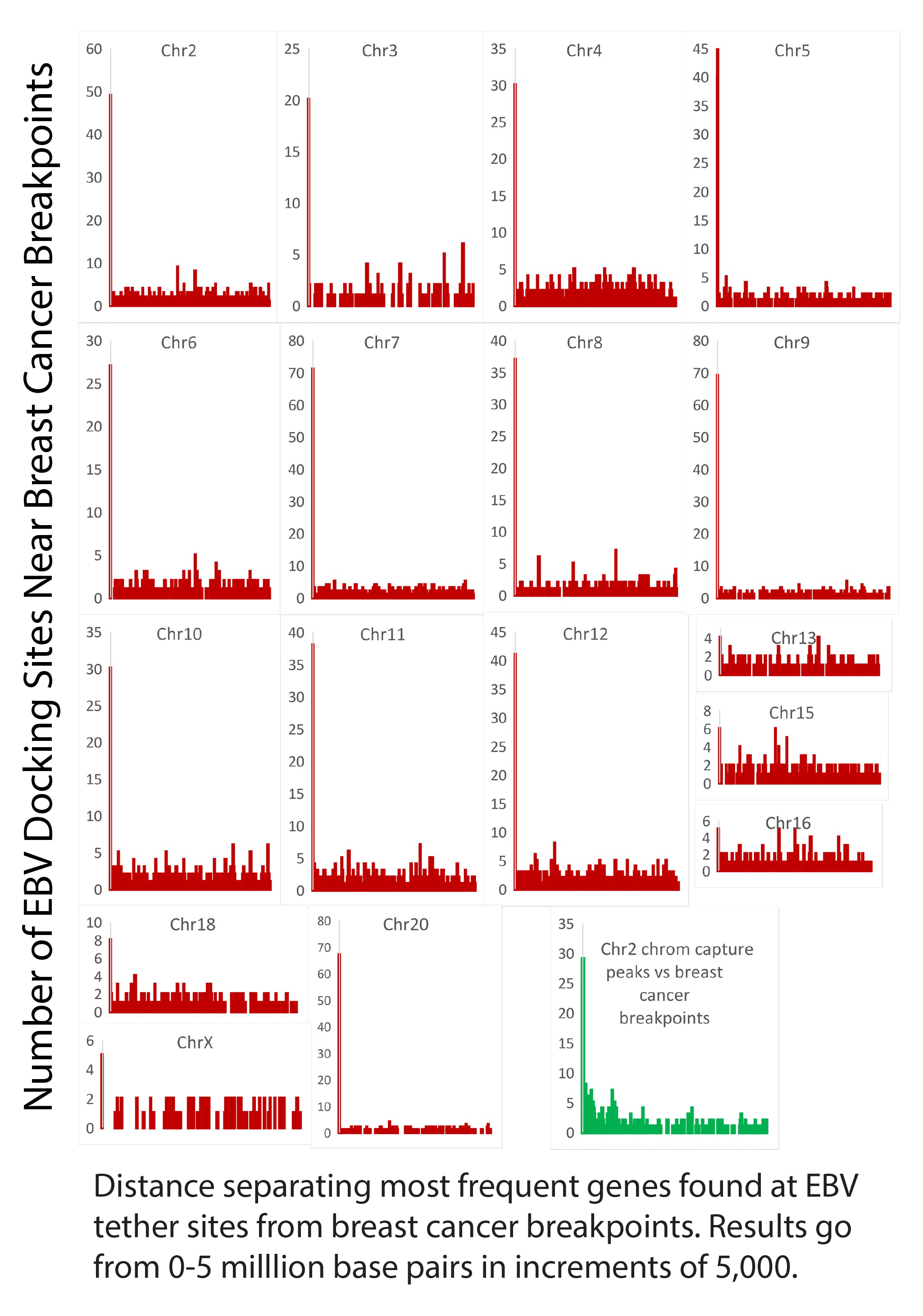
EBV anchor sites on chromosome 2 in EBV-positive cells are close to breast cancer breakpoints. Breast cancer breakpoints cluster around **t**he top 10% most frequently found genes near EBV tethering sites in BL cells.

Much additional evidence supports EBV docking to all chromosomes in cancers. Earlier results produce 903 EBNA1 binding sequences scattered over all human chromosomes (*78*). Forty fragile sites strongly bind EBNA1 at 168 positions in the human genome that do not even include chromosome 11 (*80*). X-ray crystallography of EBNA1 bound to an 18-base pair palindrome requires very high concentrations of two fixed sequences. Fig 3B shows that EBNA1 coding sequences are unlikely to be fixed in tumor variants, so they are unlikely to dock at a constant locus.

The simplest explanation for the relationship between EBNA1 binding and breast cancer breakpoints is that EBNA1 binding generates a consistent pattern of chromosome breakpoints near where it attaches and then interferes with breakpoint repair. The binding of EBNA1 sequence variants drives EBV lytic gene expression and increases the risk for NPC (*81*). Lytic genes include EBV-encoded nucleases BGLF5 (*82*) and BALF3 (*83*). EBV-related tumor viruses need these nucleases for lytic replication (*84*).

The next question was whether EBV or other viruses then cause clustered rearrangements and mutation storms. Fig. 5C shows clustered rearrangements on chromosome 6 (*42*). One thousand ninety genome coordinates describe fragments with copy number >=3. These coordinates are unlikely to be random since they do not follow a normal distribution (p<0.0001). Viral sequence coordinates strongly correlate by simple linear regression analysis (r2=0.93). This correlation suggests that repetitive virus sequences confuse repairs and contribute to clustered rearrangements.

**Fig. 5C.**
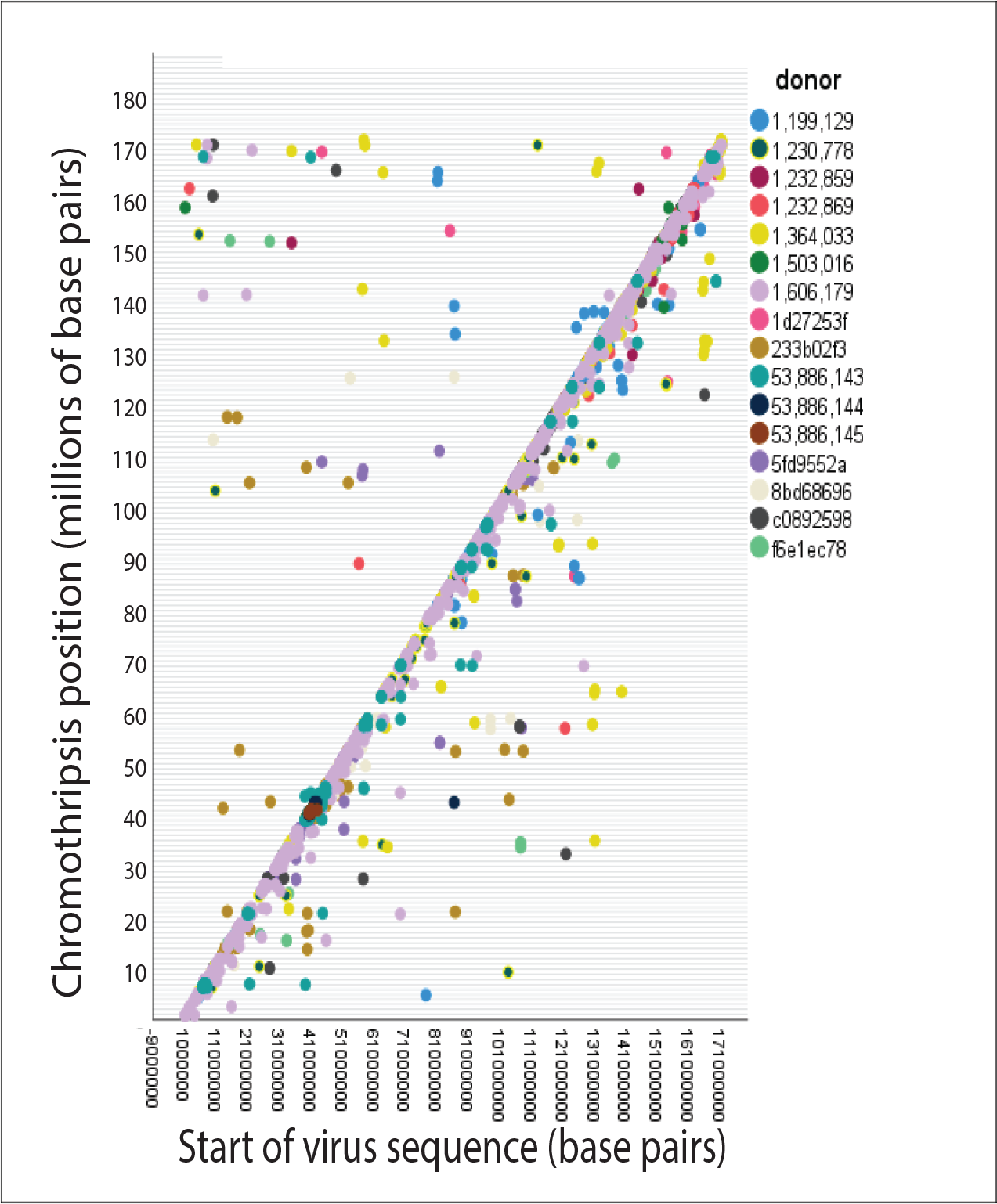
High-confidence positions where chromosome 6 shatters in 16 breast cancer genomes (*42*) plotted against start points of viral sequence homologies.

Finally, I determined whether the ability to cause DNA breaks is a characteristic property of herpesviruses. EBV has significant homology to many Kaposi’s Sarcoma virus (KSHV/HHV-8) isolates (Fig. 5D). KSHV infection causes DNA breaks (*85*) as it hijacks DNA repair proteins to favor KSHV replication (*86*). Chromosomes in infected cells are unstable, misaligned, and form anaphase bridges (*87*). Cytomegalovirus (CMV) and herpes simplex 1 (HSV1) are more distantly related to EBV but have similar structures. HCMV and HSV1 both break DNA and fragment chromosomes (*88*).

**Fig. 5D.**
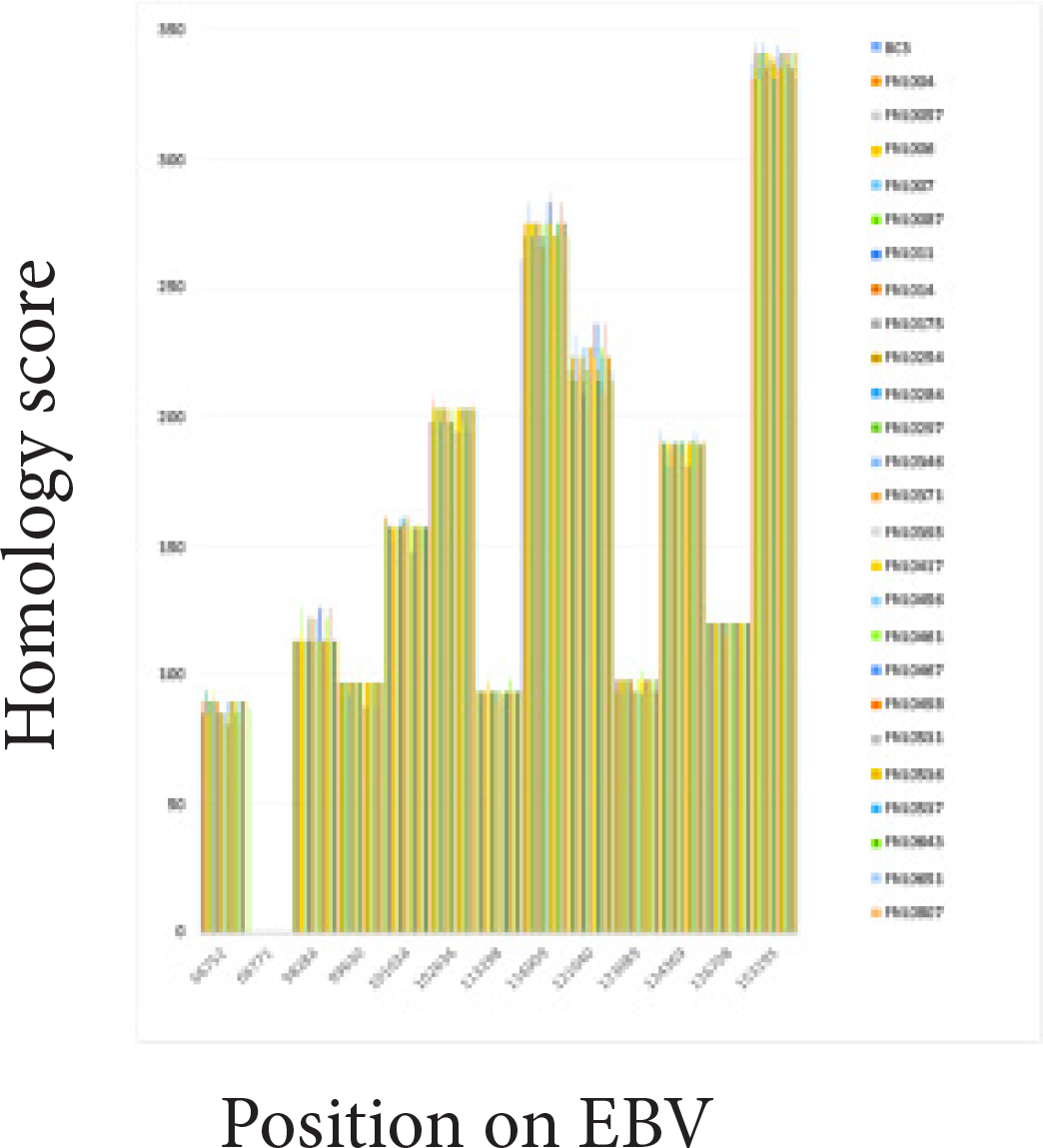
EBV is homologous to KSHV (herpesvirus 8), which is known to cause chromosome breaks.

### Alternate explanations for breast cancer breakpoints that do not involve EBV variants

Testing alternate explanations for what causes breast cancer breaks now becomes essential. Mutagens that are not viruses can cause random DNA breaks. However, elaborate systems repair this damage. Random breakpoints are not predominant in breast cancer. Breakpoints on any chromosome are unlikely to fit a normal distribution (TableS1, p<0.0001).

Tumor-infiltrating lymphocytes (TILs) are biomarkers for predicting breast cancer prognosis (*89, 90*). To test whether TILs cause chromosome breaks, I compared breakpoint numbers in 16 breast cancers with severe lymphocyte infiltration vs. 17 breast cancers with nil lymphocyte infiltration. The student’s t-test could not reject the null hypothesis that the numbers of breakpoints were statistically identical (p=0.7) (Fig. 5E).

**Fig. 5E.**
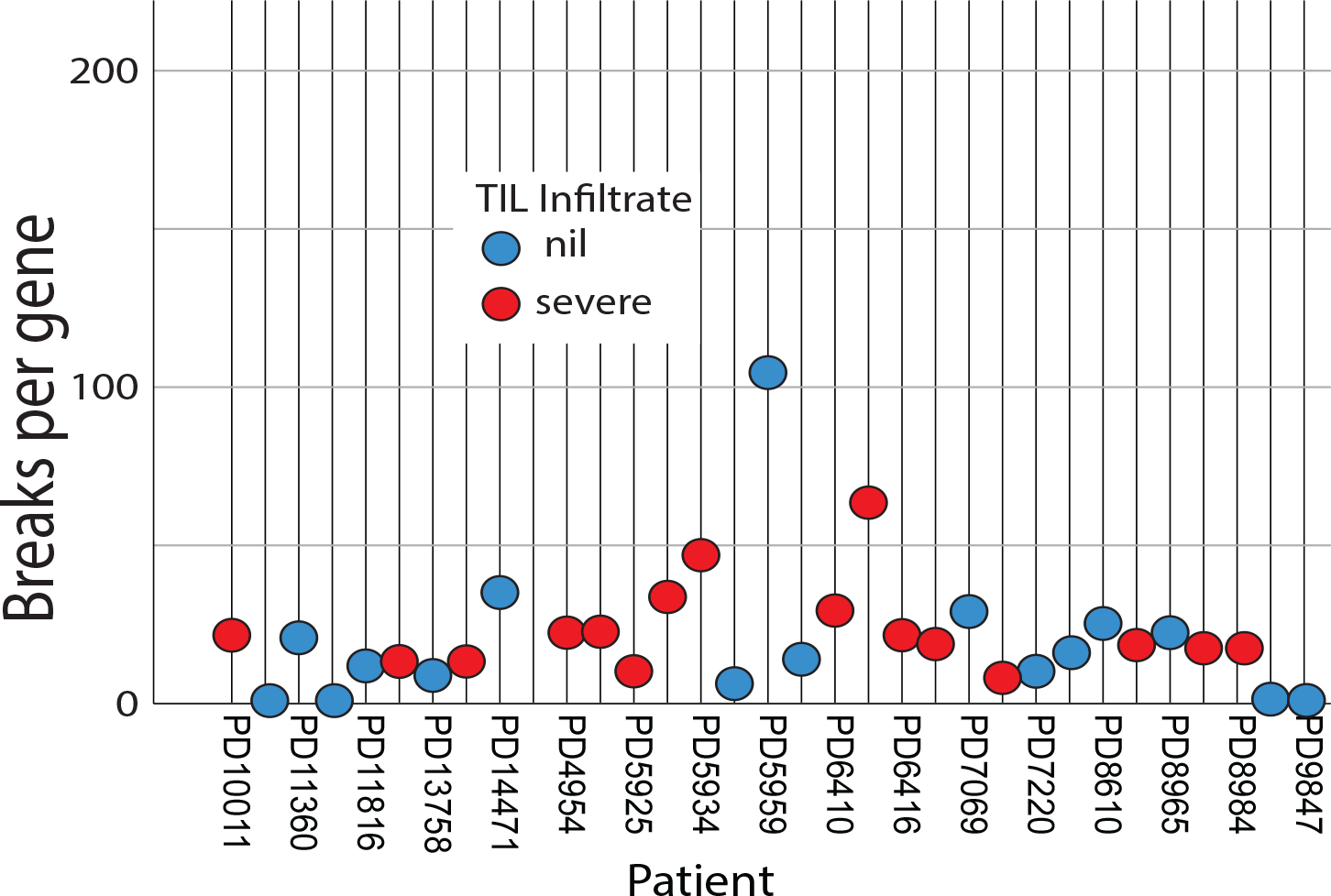
Data do not establish an impact of severe vs. nil lymphocyte infiltration (TIL) on breast cancer breakpoints.

I estimated retrovirus contributions to structural variations using data from cancer in 38 different tissues (89). Retrotransposons make relatively modest contributions to breast cancer compared to, say, esophageal or oral (gums) cancer (Fig. 5F). Retroviral sequences constitute ∼8% of the human genome. EBV can trans-activate endogenous retroviruses (*19, 91, 92*). DNA near some breast cancer breakpoints resembles porcine endogenous retrovirus (PERV), human endogenous retrovirus (HERV), and HIV1 (e.g., Figs. 3A). Breast cancers contain retrovirus MMTV (mouse mammary tumor) DNA (*93, 94*). The human reference genome has MMTV provirus sequences at 24 places. Matches (bit scores 854-1164) include a retrotransposon, an envelope protein, and an LTR. Envelope pseudogenes give another ten scores of ∼300. HPV variants are DNA viruses also implicated in breast cancer. HPVs were not assessed further, but they occasionally match DNA near breast cancer breakpoints.

**Fig. 5F.**
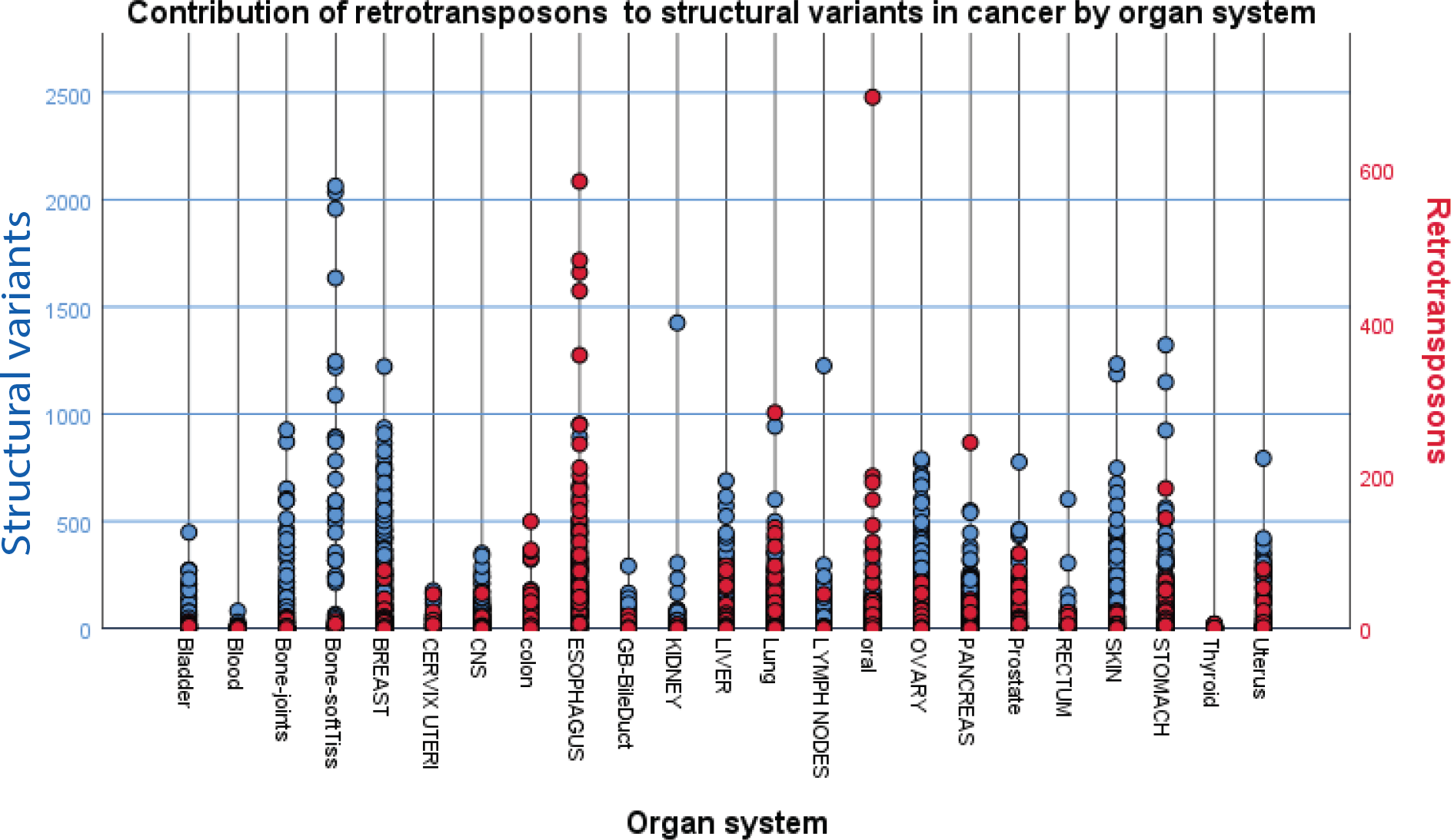
Retrotransposons make relatively minor contributions to breast cancer structural variants compared to cancers at other sites.

Common fragile sites are site-specific breaks seen on metaphase chromosomes after inhibiting DNA synthesis by DNA polymerase inhibitors. Some common fragile sites (*95*) align with breast cancer breaks on chromosome 1, but breakpoints on most other chromosomes are incompatible. Chromosomes 8, 9, 11-15,17-19, 21, and 22 do not have common fragile sites but still have many breast cancer breaks (*47*).

### EBV and Metastasis

At this stage, the work has found links between activated EBV and breast cancer. So I asked whether EBV contributes to breast cancer metastasis. According to Yates and coworkers, relapsed and metastatic tumors keep their tumor-driver gene mutations and continue acquiring new ones. Eventually, late mutations in JAK-STAT and SWI-SNF signaling drive established breast cancers into metastasis (*96*). Forty-five percent of breast cancers in COSMIC have mutations in JAK and STAT isoforms. The other Yates metastasis-shifting mutations affect SWI-SNF (*96*), a complex that repositions nucleosomes and supports genome stability (*97*). The FA-BRCA pathway senses obstacles to replication, and SWI-SNF addresses them (*97, 98*). Breast cancers consistently disable SWI-SNF by targeting ARID genes (*96*). ARID1A is a COSMIC top 20 most frequently mutated gene in breast cancer.

EBV facilitates its replication by disabling JAK-STAT and SWI-SNF signals, the same losses that shift breast cancer into metastasis. First, the EBV immune evasion protein BCRF1 (viral IL-10) binds to interferon receptors to prevent triggering JAK-STAT-mediated immune responses (*99*). Meanwhile, the EBV tegument protein BGLF2 suppresses JAK-STAT signaling. BGLF2 further prevents interferon-stimulated gene activation (*100*). EBV also produces an adapter protein (EBNA2) that inhibits the SWI-SNF complex (*101*). EBNA2 binding to SWI-SNF cripples homologous recombination.

Mutations in EBV cancers such as NPC prove that Yates metastasis driver gene damage accompanies EBV infection. NPC often loses Type 1 interferon genes (IFNA1, IFNA2, IFNA8, and IFNE) and nearby MTAP (34%) by homozygous deletions at chr9p21.3 (*19*). This deletion prevents JAK-STAT signaling. Breast cancers (TableS1) have 65 breakpoints strictly within this interferon-MTAP region (chr9:21579478-20503534), not counting longer fragments that include the interval. Like breast cancer, NPC has multiple recurrent aberrations in ARID1A and ARID1B, which disable SWI-SNF. NPC often inactivates SWI-SNF components BAP1 and PBRM1 within a frequently damaged 3p21.3 gene cluster (*19*) at chr3:52400000-53000000. Breast cancers have 18 breakpoints within this short interval. DLBCL, another EBV-linked cancer, has a dampened interferon response (*102*) and recurrent alterations in SWI-SNF complexes (*103*).

## Discussion

Human papillomavirus promotes cervical cancer (*2*) because it disables tumor suppressors. EBV in breast cancer resembles this model. EBV variants disable a variety of molecular and cellular safeguards that protect against breast and other cancers. EBV-mediated cancers have deficits in the tumor suppressor FA-BRCA pathway (*19, 22*), the same pathway that underlies hereditary breast and ovarian cancer. EBV variant activations become equivalent to inheriting a high-risk BRCA1 or BRCA2 mutation. EBV even produces a micro-RNA that inhibits BRCA1 (*23, 24*). Further, the virus degrades SMC5/6 (*26*), a repair scaffold interacting with critical FA-BRCA intermediates. EBV also causes the same gene losses that permit established breast cancers to become metastatic. A summary of multiple independent lines of evidence supports this view (Fig. 6A).

1. Breakpoints in BRCA mutation-positive and negative breast and ovarian cancers cluster near breakpoints in diverse EBV-related cancers such as NPC, DLBCL, and BL. Cancers not related to EBV do not have this clustering.
2. Breaks in HER2-positive and triple-negative breast cancers target chromosomes essential for immunity, facilitating viral infections.
3. Compromised DNA repair and amplified centrosomes do not require the continuing presence of active viruses and do not depend on their levels.
4. Chr6 in breast cancer has an EBV infection signature, direct evidence for EBV infection. An EBV methylation signature shared with known EBV-related cancers affects a genome area encoding MHC genes. The signature is far more abundant in 1538 breast cancers than normal controls.
5. piRNA sequences make viral sequences into sandwiches. This sandwiching spans MHC class I, II, and III antigen DNA on chromosome 6, interspersed with the methylation signature for EBV infection. The sandwiches are presumptive evidence of past infection.
6. Breast cancer breakpoints cluster around EBV docking sites.
7. EBV disables accurate chromosome repairs and creates rearrangement clusters, multiple centromeres, and mutation storms.
8. EBV suppresses JAK-STAT and SWI-SNF signaling. This suppression facilitates EBV replication. Damage to these signals pushes breast cancer into metastasis.
9. The results rule out random breakpoint occurrences and fragile site sequences.

**Fig. 6A.**
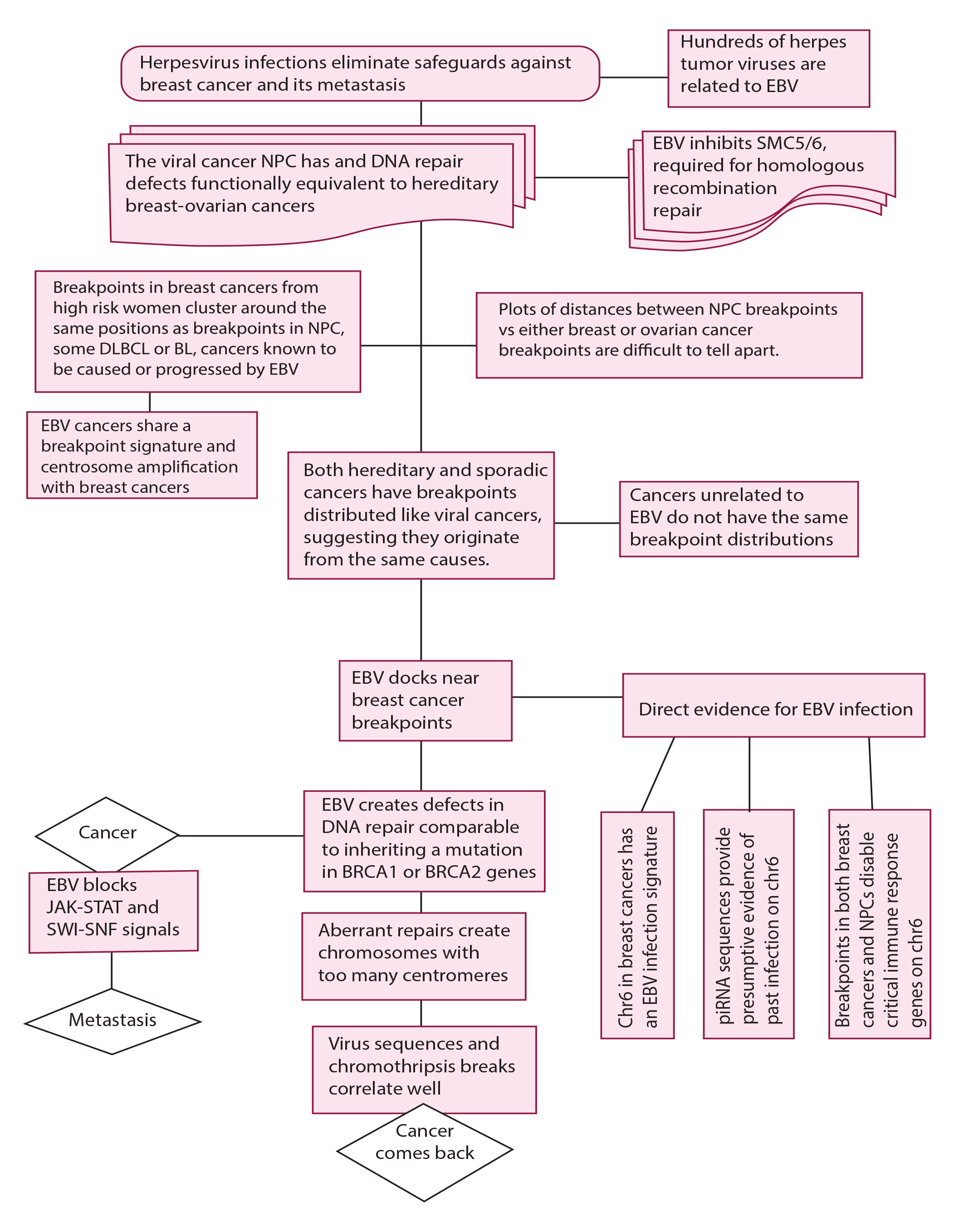
Flow chart summarizing the evidence for the model presented in Fig 7B.

### EBV infection is comparable to inheriting a breast cancer pathogenic mutation (Fig. 6B)

**Fig. 6B.**
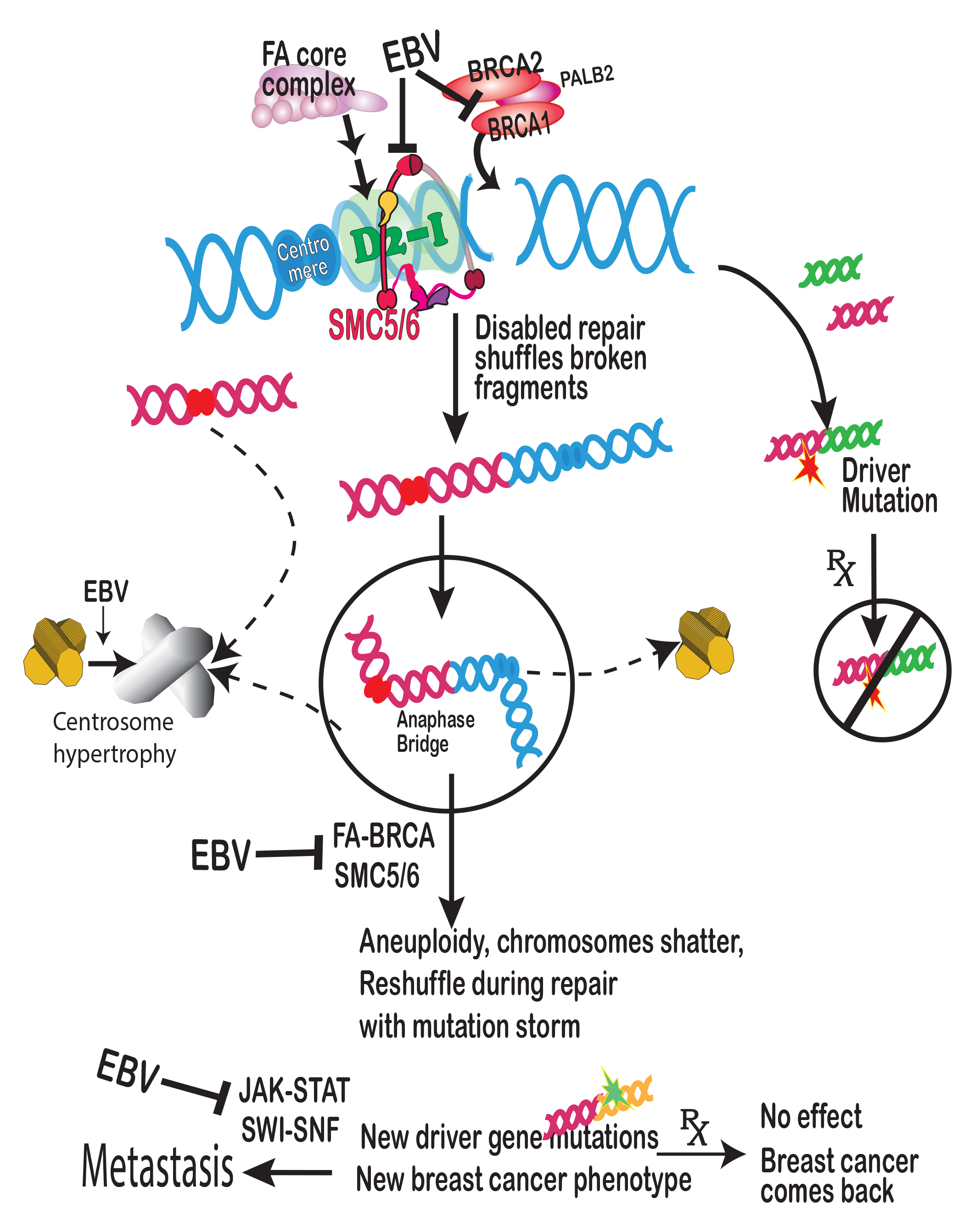
Proposed model explaining results.

A damaged immune system is characteristic of breast cancers (*104, 105*) and EBV cancers. EBV cancers share other deficits with breast cancers. The shared deficits interfere with restoring the genome from damage by natural processes and exogenous mutagens (*106*). This damage creates DNA crosslinks and DNA double-strand breaks. If crosslinks and DNA breaks persist during cell division, they cause chromosome rearrangements and cancer. Humans devote a significant resource to repairing these lesions because they threaten survival. This resource is a sprawling, interconnected repair system. The system encompasses the BRCA pathway, FA proteins, an SMC5/6 scaffold, JAK-STAT signaling, and the SWI-SNF chromatin remodeling complex.

Breast cancer breakpoints cluster around EBV binding sites, suggesting EBV participates in causing the breaks. The breaks lead to pathogenic chromosome rearrangements because EBV forces repairs by error prone methods. These error prone methods predominate when EBV suppresses key intermediates in the FA-BRCA pathway (*23, 24*). The mechanisms of NPC and breast-ovarian cancer share another deficiency because FA-BRCA-dependent repairs need access to crosslinked and broken chromosomes. Chromatin access requires the SMC5/6 cohesin complex (*28, 29*). SMC5/6 promotes sister chromatid recombination, the same process that requires BRCA1 and BRCA2 (*107*). In one scenario (Fig. 6B), SMC5/6 interacts with a crucial intermediate in the FA-BRCA pathway, the FANCD2-FANCI heterodimer ("D2-I" in Fig. 6B)(*26, 108*). If EBV variants undergo lytic activation, they degrade and deplete SMC5/6. Scarce SMC5/6 further prevents immune responses to EBV (*27*). Collateral damage is that SMC5/6 cannot facilitate FA-BRCA-mediated DNA repairs or interact with FANCD2-FANCI. In this way, EBV inhibits removing DNA crosslinks, encourages double-strand breaks and shuffles broken DNA fragments. EBV activation is like a mutagen that disables BRCA1 and BRCA2 (Fig. 6B).

EBV infection also equates to JAK-STAT and SWI-SNF signaling mutations that Yates and coworkers (*96*) report push breast cancer into metastasis. The viral immune evasion protein BCRF1 suppresses interferon responses (*99*). EBV tegument protein BGLF2 promotes STAT2 proteolysis, suppressing JAK-STAT signaling (*100*). BGLF2 prevents the expression of interferon-stimulated genes. EBV nuclear antigen (EBNA2) associates with SWI-SNF to hijack the complex to viral and cellular genes (*101*). BRG1, the human SWI/SNF chromatin modifier, supports virus transactivation of late promoters (*109*). EBV infection in NPC interferes with JAK-STAT and SWI-SNF functions (*19*)

Breast cancers in the present study are primary stage II or III cancers. Although surgery usually removes these tumors, cancer cells can transition to metastases or local recurrence. The transition requires a well-established tumor. This delay in cancer dissemination makes primary tumors accurately represent metastatic or locally relapsed cancers, so primary tumors are valid to predict metastasis and make treatment decisions (*96*).

NPC is unlikely to require inheriting BRCA1 and BRCA2 mutations. The genes are not identified as NPC-susceptibility genes (*110, 111*). Breakpoints in sporadic breast cancer and NPC are similar. Inhaled aldehyde carcinogens (e.g., formaldehyde) that expose the nasopharynx can degrade BRCA2 (*112, 113*). Many metastatic breast cancers with deficits in homologous recombination-mediated repair do not have inherited BRCA1 and BRCA2 mutations (*114*).

Breast cancers need not contain large numbers of viral particles because they are not required to provoke cancer. EBV-infected B-cells and breast cancer cells both have amplified centrosomes. Overduplication of these mitosis-organizing centers causes errors in chromosome segregation and then genomic instability (*45, 46*). FA-BRCA deficient repair of DNA breaks causes too many centromeres. When mitosis pulls apart multi-centromere structures, the forces pulverize the chromosome and induce mutation storms (*42*)(Fig. 6B). Neither centrosome amplification nor chromosome fragmentation (chromothripsis) requires large numbers of viral particles or active infection.

Recent cancer drug therapy focuses on finding and targeting cancer driver mutations. The drugs are initially effective, sometimes for long periods, but then stop working. The cycles in Fig. 6B are an occult, underlying process that continues even if cells with the initial driver mutations are removed. Cancer treatment generates new clones that do not exist in the original population (*115*). The underlying infections continually produce new cancer driver mutations.

The evidence in the present work supports therapy that considers viral infections and cells with chromosomes having multiple centrosomes and centromeres. If the ability to disable tumor suppressors and push cells into metastasis are typical properties of tumor viruses, then other viruses also contribute to breast cancer. An EBV vaccine is feasible, but the best targets are still unclear. Experiments are needed to define and characterize interactions between viruses and tumor suppressor pathways. The current evidence suggests that these experiments will be invaluable in designing a childhood herpes vaccine to prevent breast and other cancers.

## Materials and Methods

### Justification of breast cancer data choices

A primary goal of this study was to gain insight into why breast cancer occurs and recurs. Breast cancer is heterogenous, with varying levels of chromosome instability in different subtypes. To understand the underlying origins of breast cancer genomic instability, published and curated breast cancer data was the starting point. This publicly available DNA sequence data was chosen to encompass diverse genetics, subtypes, stages, grades, morphologies, and outcomes. Initially, breast cancers were separated only broadly into those with a likely hereditary component vs. those without this component. Although the selected cancers are not a representative random sample of all breast cancers (*116*), they are nonetheless likely to present with chromosome instability originating from diverse typical causes.

### Inclusion criteria for breast cancer data

Criteria for including breast cancer data required published chromosome intra-chain or inter-chain chromosome breakpoints from high-quality peer-reviewed publications produced by world-class laboratories. A second criterion was the availability of DNA sequence data sufficient to specify the location of these chromosome breakpoints. The third criterion was that genome sequencing was done on samples taken before treatment began. The cancers had to include typical morphologies such as ductal carcinomas, lobular carcinomas, medullary carcinoma, and invasive carcinoma, "no special type. The samples were all primary tumors at different stages, or in one case, a primary tumor reappearing locally.

### Breast cancers in high-risk women

Hereditary cancers were taken as breast cancers from women with a typed high-risk BRCA1 or BRCA2 mutation diagnosed before age 70. To obtain more data, cancers with onset before age 50 were also included since these are at high risk for an inherited cancer-associated mutation.

### Sporadic breast cancers

Sporadic breast cancers are those diagnosed after age 70 without a known inherited mutation.

### Original data sources for breast cancer data

The raw data comes from 560 breast cancer genome sequences, familial cancer data from 78 patients, methylation data from 1538 breast cancers vs. 244 controls, 243 triple-negative breast cancers, and 2658 human cancers (*13, 42, 52, 54, 117*). In all, seventy-four breast cancers from high-risk women were typed BRCA1 or BRCA2 mutation-associated or cancers diagnosed before age 40 (*118, 119*). Another study of familial breast cancers contributed sixty-five familial breast cancers (*54*). HER2-positive and triple-negative breast cancer data was from original publications (*54*) and the COSMIC website. Gene breakpoints for inter-chromosomal and intra-chromosomal translocations and breakpoints were obtained from the COSMIC website as curated from original publications or from original articles and their supplemental information (*52, 54, 117*).

### Original data sources for NPC, DLBCL, and BL cancers with known EBV associations

NPC chromosome breakpoint positions are from Bruce et al. (*19*) for 70 primary tumors of the nasopharynx at stages 1-IVC. The data come from whole-genome sequencing of "63 micro-dissected tumors, 5-patient derived xenografts, and two cell lines". DLBCL breakpoints were from 88 DLBCL patients (age >60y) (*37*). The MYC breakpoints include class I and II MYC translocation breakpoints defined in BL, encompassing areas far upstream of c-myc (*120, 121*). Downstream breakpoints included an enhancer region about 565 kb on the telomere side of the MYC coding sequence (*37*). BL breakpoints (*66*) were converted to GRCh38/hg38. Older data gives fusion sequences as Gencode Accession numbers (*35*). Fusion sequences were downloaded as a FASTA file and copied to BLAT for placement on the human GRCh38/hg38 reference sequence,

### Data source for ovarian cancers

Data for breakpoints in ovarian cancers were downloaded from the COSMIC website. The cancers are "mixed adenosquamous ovarian carcinomas" and are arbitrarily taken from those with the largest number of structural variants. Cancers all had the prefix AOCS-with further identification numbers and BRCA mutation status as follows: 170-1-8 (neg), 120-3- 6(BRCA2), 142-3-5 (neg), 139-1-5 (neg), 086-3-2 (neg), 147-1-1 (BRCA1, BRCA2), 094-6-X (BRCA1), 094-1-1 (BRCA1), 088-3-8 (neg), 139-6-3 (BRCA2), 150-3-1(neg), 116-1-3 (neg), 155-3-5 (BRCA2), 093-3-6 (neg), 034-3-8 (BRCA1), 091-3-0 (BRCA1), 139-19-0 (BRCA2), 170-3-5 (neg), 114-1-8 (neg), 064-3-3 (neg), 064-1-6 (neg), 106-1-1 (BRCA1), 152-1-X (BRCA1), 134-1-5 (?).

### Exclusions

Male breast cancers were excluded.

### Comparisons of DNA sequences

The UCSC online "Liftover" function interconverted different versions of genome coordinates into GRCh38/hg38 coordinates, such as those for breakpoints, viral-human similarities, and piRNA locations. DNA flanking sequences at breakpoints were downloaded primarily from the GRCh38/hg38 version of the UCSC genome browser as FASTA files and inputted directly into BLAST. Results were checked against breakpoints in 101 triple-negative breast cancers from a population-based study (*117*). The GRCh38/hg38 human genome version was used whenever possible, but some comparisons of chromothripsis breakpoints were more straightforward with earlier versions. piRNA locations were from the piRNA bank (*122, 123*). Positions of differentially methylated regions near breast cancer breakpoints (*11*) were compared to breakpoint positions for 70 NPCs based on published data (*19*).

### Calculation of distances between breakpoints in different cancers

The nearest break position in breast cancer to a break in NPC was taken as the XLOOKUP value for the number of base pairs from the closest NPC breakpoint 5’ to the breast cancer break or the NPC breakpoint 3’ to the breast cancer break, whichever was closer (supplementary TableS1). Distance from the breast cancer breakpoint was then calculated as the absolute value of the difference between the closest NPC break and the tested breast or ovarian cancer break. Differences in the amount of data available for NPC vs. breast cancer breakpoints complicated the calculations near the ends of chromosomes. Several methods of handling these end regions made no discernible difference in the outcomes. For a 5,000 base pair window, an overflow window of 5,000,000 was used to limit the number of bins to a maximum of 1,000. The validity of the calculations was further tested for a few chromosomes by scanning the breast cancer breakpoints vs. the NPC breakpoints in the reverse direction. The same results were obtained. Millions of calculations were repeated at least twice.

### DNA sequence homologies

The NCBI BLASTn program (MegaBLAST) and database (*124-126*) compared DNA sequences around breakpoints in breast cancers to all available viral DNA sequences. E(expect) values are related to p values and represent the probability that a given homology bit score occurs by chance. E values <1e-10 were considered significant homology. In many cases, E values were "0" (<1e-180) and always far below 1e-10. Virus DNA was from BLAST searches using "viruses (taxid:10239)" with human sequences and uncharacterized sample mixtures excluded. Different strains and isolates of the same virus were tested for human homology. Specifically, HKHD40 and HKNPC60 were often considered together as "EBV." Homology between these virus groupings vs. human sequences was determined using "Blastn." Human and mouse sequences were excluded.

### EBV binding site locations and palindromes

EBNA1 binding locations (*78*) specified genes within or near EBV DNA binding locations. Breaks in breast cancers were compared to the gene positions around their EBNA1 binding sites. The Palindrome Site Finder from NovoPro and EMBOSS palindrome identified palindromic DNA sequences.

### Comparisons among human herpes viruses

EBV variants HKHD40 and HKNPC60 were compared to human herpes viruses in BLASTn by entering the terms human gamma herpesvirus 4, herpesviridae, and herpesvirales. I used values with >= 2000 base pairs in common. I also tested the EBV reference sequence against known cancer viruses HHV8, HSV1, and HCMV.

### Data analyses

Microsoft Excel, SPSS, StatsDirect, Visual Basic, and Python + Biopython (*127*) scripts provided data analysis. The NCBI Genome Decoration page and the Ritchie lab website (*128*) provided chromosome annotation software.

### Statistics

Statistical analyses used StatsDirect, SPSS statistical software, or Excel. A scatter plot of the data was always prepared before running any statistical analysis. The Fisher exact test compared viral similarities around breakpoints in hereditary breast cancers to similarities in breakpoint positions generated by random numbers. Unpaired t-tests and Mann-Whitney U tests compared breakpoint distributions. Tests for normality included kurtosis and skewness values and evaluation by methods of Shapiro-Francia and Shapiro-Wilk (*129*). Details are included with the breakpoint calculations in supplementary TableS1.

### Fragile site sequence data

Positions of fragile sites were from a database (*95*) and original publications (*130*).

### Data Availability Statement

The primary datasets generated or analyzed during this study are included in the published article and its supplementary information files. Datasets not included are freely available from the corresponding author on reasonable request.

